# SARS-CoV-2 infections during Omicron (BA.1) dominant wave and subsequent population immunity in Gauteng, South Africa

**DOI:** 10.1101/2022.07.13.22277575

**Authors:** Shabir A. Madhi, Gaurav Kwatra, Jonathan E. Myers, Waasila Jassat, Nisha Dhar, Christian K. Mukendi, Lucille Blumberg, Richard Welch, Alane Izu, Portia C. Mutevedzi

**Author notes:** **Corresponding author:** Shabir A. Madhi, University of the Witwatersrand, Phillip Tobias Building., Princess of Wales St., Parktown, 2193, Gauteng, South Africa. Equal contribution to the study (Gaurav Kwatra, Alane Izu, Portia C. Mutevedzi).

## Abstract

**Background:** The B.1.1.529 (Omicron BA.1) variant of severe acute respiratory syndrome coronavirus 2 (SARS-CoV-2) caused a global resurgence of coronavirus disease 2019 (Covid-19). The contribution of BA.1 infection to population immunity and its effect on subsequent resurgence of B.1.1.529 sub-lineages warrant investigation.

**Methods:** We conducted an epidemiologic survey to determine the sero-prevalence of SARS-CoV-2 IgG from March 1 to April 11, 2022, after the BA.1-dominant wave had subsided in Gauteng (South Africa), and prior to a resurgence of Covid-19 dominated by the BA.4 and BA.5 (BA.4/BA.5) sub-lineages. Population-based sampling included households in an earlier survey from October 22 to December 9, 2021 preceding the BA.1 dominant wave. Dried-blood-spot samples were quantitatively tested for IgG against SARS-CoV-2 spike protein and nucleocapsid protein. Epidemiologic trends in Gauteng for cases, hospitalizations, recorded deaths, and excess deaths were evaluated from the inception of the pandemic to the onset of the BA.1 dominant wave (pre-BA.1), during the BA.1 dominant wave, and for the BA.4/BA.5 dominant wave through June 6, 2022.

**Results:** The 7510 participants included 2420 with paired samples from the earlier survey. Despite only 26.7% (1995/7470) of individuals having received a Covid-19 vaccine, the overall sero-prevalence was 90.9% (95% confidence interval [CI], 90.2 to 91.5), including 89.5% in Covid-19 unvaccinated individuals. Sixty-four percent (95%CI, 61.8-65.9) of individuals with paired samples had serological evidence of SARS-CoV-2 infection during the BA.1 dominant wave. Of all cumulative recorded hospitalisations and deaths, 14.1% and 5.9% were contributed by the BA.1 dominant wave, and 5.1% and 1.6% by the BA.4/BA.5 dominant wave. The SARS-CoV-2 infection fatality risk was lower in the BA.1 compared with pre-BA.1 waves for recorded deaths (0.02% vs. 0.33%) and Covid-19 attributable deaths based on excess mortality estimates (0.03% vs. 0.67%).

**Conclusions:** Gauteng province experienced high levels of infections in the BA.1 -dominant wave against a backdrop of high (73%) sero-prevalence. Covid-19 hospitalizations and deaths were further decoupled from infections during BA.4/BA.5 dominant wave than that observed during the BA.1 dominant wave.

(Funded by the Bill and Melinda Gates Foundation.)

## BACKGROUND

As of November 14, 2021, an estimated 44% of the world population was infected at least once by severe acute respiratory syndrome coronavirus-2 (SARS-CoV-2) since the start of the coronavirus disease (Covid-19) pandemic in March 2020^1^. In mid-November 2021, the highly transmissible Omicron BA.1 (B.1.1.529; henceforth referred to as BA.1) variant of concern (VOC) was identified in Southern Africa, heralding a resurgence of Covid-19 globally^2^. The intrinsic transmissibility of BA.1 is estimated to be twice as high as that of the Delta VOC which has a basic reproduction rate (*Ro*) of 5-6^3^. The BA.1 VOC is resistant to neutralizing activity of antibody induced by first-generation Covid-19 vaccines and infection by wild-type virus or earlier VOCs^4^. Nevertheless, CD4^+^ and CD8^+^ T-cell immunity against BA.1 induced by prior infection with SARS-CoV-2 ancestry virus and other variants or by Covid-19 vaccines was relatively conserved. BA.1 was associated with high numbers of infections, re-infections and breakthrough Covid-19 in vaccinated individuals^5^, which were decoupled from severe disease, probably due to underlying cell mediated immunity^6^.

Approximately three months after the BA.1 dominant wave had subsided in South Africa in April 2022, another resurgence dominated by BA.4 and BA.5 sub-lineage varinats (herewith referred to as BA.4/BA.5) which manifested relative resistance to neutralizing activity of antibody induced by BA.1 infection, more so in the unvaccinated^7^. As of mid-June 2022, BA.4/BA.5 infections are increasing in the Northern Hemisphere, and are anticipated to become dominant in the European Union in the near future^8^.

Our previous population based sero-survey in Gauteng Province, South Africa, reported 73% of the population had acquired SARS-CoV-2 immunity prior to the onset of the BA.1 dominant wave despite only 19% having received a single dose of Covid-19 vaccine^9^. Extensive infection-induced cell mediated immunity likely contributed to the decoupling of infection and severe disease observed during the BA.1 dominant wave.

In this study, we report on a follow-up sero-survey conducted after the BA-1 dominant wave had subsided, and which coincided with the onset of the BA.4/BA.5 dominant wave. Using paired serum anti-nucleocapsid (anti-N) and anti-spike (anti-S) IgG responses from individuals included in the current and preceding survey, population rates of serologically identified SARS-CoV-2 infections during the BA.1 dominant wave were inferred. We updated incidence rates of cases, hospitalizations and deaths including the current BA.4/BA.5 dominant wave in Gauteng. Serological and epidemiological data allowed estimation of ratios of inferred infections to reported cases, hospitalizations and fatalities, as well as Infection Fatality Risk (IFR) for the first three Covid-19 waves cumulatively (pre-BA.1), and the BA.1 dominant wave.

## METHODS

### Study setting and data collection

A third cross-sectional population-based sero-survey in Gauteng, South Africa was conducted between March 1^st^, 2022 and April 11^th^, 2022; Supplementary Table S1. Details of the study setting, and methods of the survey have been previously described^9,10^ and can be found in the supplementary materials.

The survey was done in partnership with Gauteng Department of Health as part of public health surveillance. The Human Research Ethics Committee at the University of the Witwatersrand granted a waiver for ethics approval of the survey.

### Serological analysis

The serology testing for anti-nucleocapsid (anti-N) and anti-spike (anti-S) IgG was done on dried blood spots samples obtained from the participants as previously described^9^, and briefly detailed in the supplementary appendix.

### COVID-19 data sources

Data sources as previously described and include daily recorded Covid-19 cases, hospitalisations and deaths to June 6^th^, 2022 from the National Institute for Communicable Diseases (NICD) in South Africa ^11^, and excess deaths attributable to Covid-19 (all excess deaths were assumed to be COVID-19 deaths) to 4 June, 2022 from the South African Medical Research Council^12^. The mid-2021 Gauteng province population projections from Statistics South Africa (STATS-SA) were used^13^.

### Statistical analysis

The anti-N IgG sensitivity for detecting past infection was previously reported as 58.0%, hence, we used anti-N or anti-S IgG positivity to characterise overall sero-prevalence^9^. Anti-S IgG positivity in individuals who received a Covid-19 vaccine (either A26.CoV2.S or BNT162.b2) could be due to vaccination or past infection.

The criteria used to determine serological evidence of SARS-CoV-2 infection during the interval between the pre-BA.1 and post-BA.1 dominant wave sero-surveys, among those individuals with paired samples from each survey, is detailed in Supplementary Table S2a-b. In individuals who had not received any Covid-19 vaccine in the interval between the two sero-surveys and were sero-negative for anti-N or anti-S IgG in the earlier study, sero-positivity for either anti-S or anti-N IgG was defined as seroconversion, respectively. For individuals who were anti-N or anti-S IgG positive at the previous survey, sero-response was defined by a two-fold or greater increase in anti-N IgG or anti-S IgG between the two time points, respectively. The calculation of serological evidence of presumed BA.1 infections was based on either seroconversion or sero-response between the survey time-points. In individuals with paired samples who received a Covid-19 vaccine in the interval between the two sero-surveys, a twofold increase in anti-N IgG or seroconversion for anti-N IgG (i.e. negative to positive) was used as serological evidence of SARS-CoV-2 infection.

The percentage sero-positivity for either anti-N or anti-S in the Covid-19 unvaccinated individuals in the survey prior to the BA.1 dominant wave, multiplied by the STATS-SA population yielded the inferred number of infections over the course of the first three Covid-19 waves prior to the BA.1. The percentage with serological evidence of infection (composite of sero-response and sero-conversion) in the Covid-19 unvaccinated individuals with paired samples multiplied by the STATS-SA^13^ population yielded the inferred number of infections during the BA.1 dominant wave. Inferred numbers of SARS-CoV-2 infections were used to calculate ratios of inferred numbers of infections to recorded Covid-19 cases, hospitalizations and deaths; and Covid-19 attributable deaths derived from excess mortality estimates. Inferred numbers of infections at the population level allowed direct calculation of IFR which are the inverse of the inferred infections: recorded (or excess mortality attributable) Covid-19 deaths ratio. Data was analysed using R v4.1.1 (Vienna, Austria) and STATA v16.1 (College Station, USA).

### Survey Ethics

The Human Research Ethics Committee at the University of the Witwatersrand granted a waiver for ethics approval of the survey, which was being done as part of public health surveillance by the Gauteng Department of Health. All participants were, however, required to provide written informed consent; and individuals within a household were free to decline participation.

## RESULTS

### Participants

We surveyed 3345 households, including 1052 (31.4%) enrolled in the previous survey. Dried blood spots were obtained from 7510 individuals, including 2420 (32.2%) with paired samples, Figure 1 and Supplementary Table S2a. Those with and without paired samples were similar demographically and for sero-prevalence; Supplementary Table S3.

**Figure 1:**
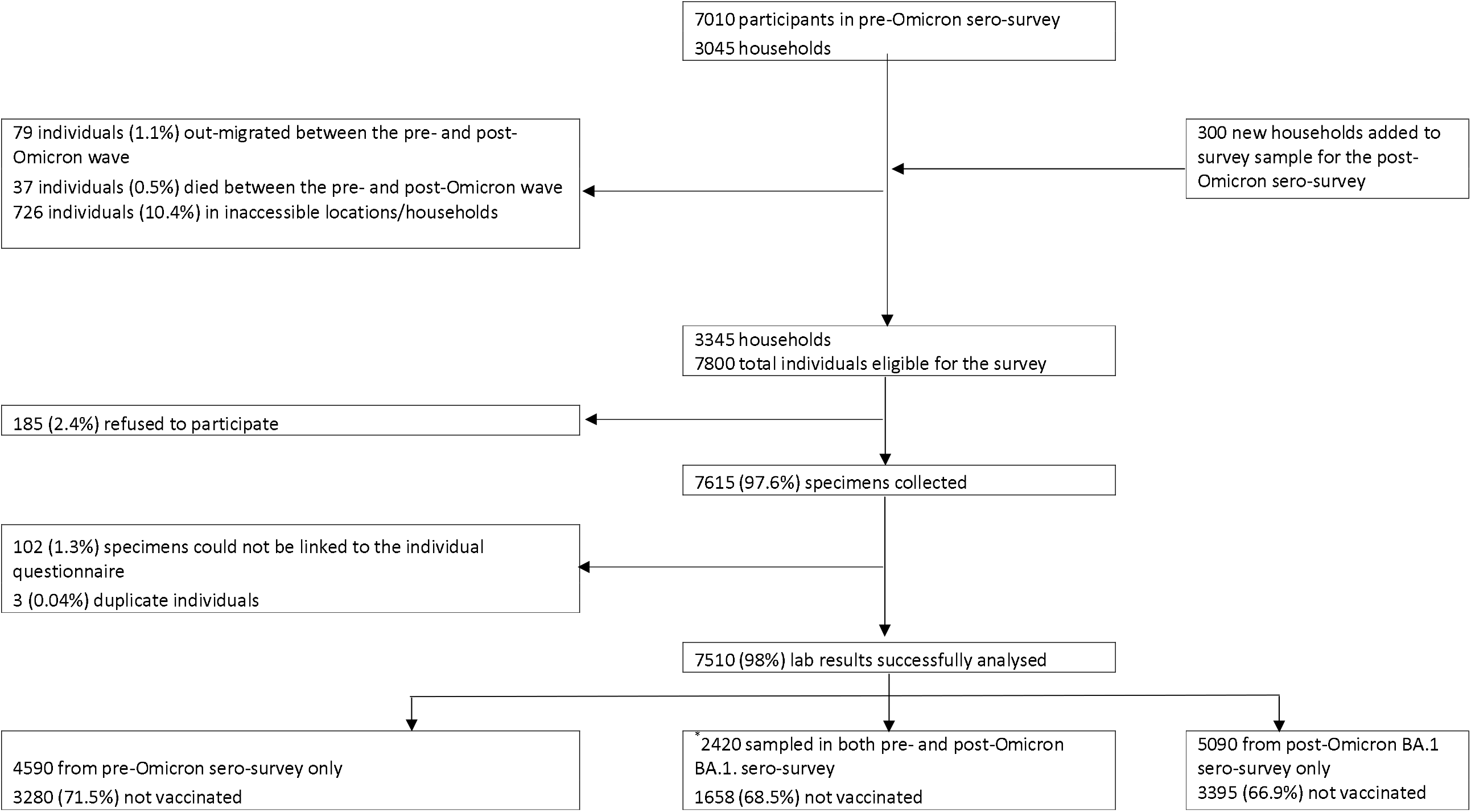
Flow of households and participants included in the seroprevalence surveys. We show flow of participants included in survey analyses from identifying the households and approaching individuals for informed consent through to specimen collection, processing, and data analyses. The final analysis included 7510 individuals in 26 sub-districts. ^*^4590 individuals from the pre-omicron sero-survey could not be sampled during the post-BA.1 dominant wave because 79 individuals (1.1%) out-migrated and 37 individuals (0.5%) died between the pre- and post-Omicron BA.1 dominant waves, 726 individuals (10.4%) could not be reached because their households were in estates where access was denied and 185 refused to participate. Finally, 3563 were unavailable for sampling in the current sero-survey.

### Seroprevalence

Current sero-positivity was higher compared with the preceding sero-survey prevalence; Supplementary Figure S1. Overall anti-S or anti-N IgG sero-positivity was 90.9% (95% confidence intervals [95%CI]: 90.2-91.5), ranging across the five districts from 86.3% to 93.0%; Table 1. Current sero-positivity was lower in the <12 year (84.1%) compared with older age-groups (>91%); Table 2.

**Table 1:** Seroconversion and changes in SARS-CoV-2 anti-spike (anti-S) or anti-nucleocapsid (anti-N) immunoglobulin G (IgG) Gauteng Province during the Omicron wave in South Africa.

| District | * Pre-BA.1 dominant wave survey |  | Post-BA.1 dominant wave survey |  | Individuals with paired samples from pre-BA.1 and post-BA.1 dominant waves sero-surveys and no Covid-19 vaccination following pre-BA.1 sero-survey<sup>1</sup> |  |  |
| --- | --- | --- | --- | --- | --- | --- | --- |
|  | N | Seroprevalence<sup>2</sup><br>n (%; 95% CI<sup>5</sup>) | N | Seroprevalence<sup>2</sup><br>n (%; 95% CI<sup>5</sup>) | Seroconversion<sup>2</sup><br>n/N (%; 95% CI<sup>5</sup>) | Seroresponse for anti-N and/or anti-S IgG<sup>3</sup><br>n/N (%; 95% CI<sup>5</sup>) | Overall serological evidence SARS-CoV-2 infection<sup>4</sup><br>n/N (%; 95% CI<sup>5</sup>) |
| **Gauteng Province** | 7010 | 5124 (73.1; 72.0-74.1) | 7510 | 6823 (90.9; 90.2-91.5) | 382/510<br>(74.9; 71.0-78.5) | 933/1548<br>(60.3; 57.8-62.7) | 1315/2058<br>(63.9; 61.8-65.9) |
| **Johannesburg District** | 2468 | 1880 (76.2; 74.5-77.8) | 2630 | 2412 (91.7; 90.6-92.7) | 124/154<br>(80.5; 73.6-86.0) | 351/574<br>(61.1; 57.1-65.1) | 475/728<br>(65.2; 61.7-68.6) |
| **Ekurhuleni District** | 1861 | 1382 (74.3; 72.2-76.2) | 2132 | 1982 (93.0; 91.8-94.0) | 133/167<br>(79.6; 72.9-85.0) | 344/529<br>(65; 60.9-69.0) | 477/696<br>(68.5; 65.0-71.9) |
| **Sedibeng District** | 564 | 398 (70.6; 66.7-74.2) | 624 | 557 (89.3; 86.6-91.5) | 21/30<br>(70; 52.1-83.3) | 44/77<br>(57.1; 46.0-67.6) | 65/107<br>(60.7; 51.3-69.5) |
| **City of Tshwane District** | 1464 | 975 (66.6; 64.1-69.0) | 1455 | 1255 (86.3; 84.4-87.9) | 72/117<br>(61.5; 52.5-69.9) | 137/269<br>(50.9; 45.0-56.8) | 209/386<br>(54.1; 49.2-59.1) |
| **West Rand** | 653 | 489 (74.9; 71.4-78.1) | 669 | 617 (92.2; 89.9-94.0) | 32/42<br>(76.2; 61.5-86.5) | 57/99<br>(57.6; 47.7-66.8) | 89/141<br>(63.1; 54.9-70.6) |
| **Age-group stratification** |  |  |  |  |  |  |  |
| **<12 years** | 753 | 423 (56.2; 52.6-59.7) | 584 | 491 (84.1; 80.9-86.8) | 53/74<br>(71.6; 60.5-80.6) | 82/126<br>(65.1; 56.4-72.8) | 135/200<br>(67.5; 60.7-73.6) |
| **12 to 17 years** | 622 | 459 (73.8; 70.2-77.1) | 553 | 523 (94.6; 92.4-96.2) | 30/33<br>(90.9; 76.4-96.9) | 94/127<br>(74; 65.8-80.9) | 124/160<br>(77.5; 70.4-83.3) |
| **18 to 50 years** | 4047 | 2978 (73.6; 72.2-74.9) | 4614 | 4204 (91.1; 90.3-91.9) | 210/270<br>(77.8; 72.4-82.3) | 495/836<br>(59.2; 55.8-62.5) | 705/1106<br>(63.7; 60.9-66.5) |
| **>50 years** | 1588 | 1264 (79.6; 77.5-81.5) | 1739 | 1587 (91.3; 89.8-92.5) | 85/128<br>(66.4; 57.9-74.0) | 256/450<br>(56.9; 52.3-61.4) | 341/578<br>(59; 54.9-62.9) |

<sup>1</sup>Of the 2420 paired samples available, 2058 did not receive a Covid-19 vaccination between the pre- and post-Omicron BA.1 sero-surveys.
<sup>2</sup>Seroprevalence was defined as seropositive for anti-S or anti-N IgG, irrespective of vaccinations status.
<sup>2</sup>Seroconversion is defined for individuals who were seronegative to both S and N at pre-Omicron sero-survey and seroconverted to either S or N at post-Omicron BA.1 sero-survey.
<sup>3</sup>Seroresponse for anti-N and/or anti-S IgG is defined for individuals who were seropositive to either S or N at pre-Omicron sero-survey and either seroconverted to S, seroconverted to N, were seropositive to N at pre-Omicron BA.1 sero-survey and had a $\geq 2$ fold increase in anti-N titers at post-BA.1 dominant wave sero-survey, or were seropositive to S at pre-BA.1 sero-survey and had a $\geq 2$ fold increase in anti-S titers at post-BA.1 sero-survey.
<sup>4</sup>Overall serological evidence of SARS-CoV-2 infection between in the period between the two surveys when the BA.1 dominant wave occurred was defined as either seroconversion or seroresponse for anti-N and/or anti-S IgG.
<sup>5</sup>CI, confidence interval; Confidence intervals have not been adjusted for multiplicity and should not be used for inference.
\* *Madhi et al, 2022*<sup>9</sup>
The criteria used for determining seroconversion and seroresponse is outlined in Supplementary Table S2b.

**Table 2:** Sero-prevalence of SARS-CoV-2 anti-spike or anti-nucleocapsid immunoglobulin G and risk factors for sero-positivity in Gauteng Province, stratified by sex, age group, and district.

| Category | Pre-BA.1 dominant wave sero-survey<sup>1</sup> |  | Post-BA.1 dominant wave sero-survey |  |
| --- | --- | --- | --- | --- |
|  | Number sampled<br>N (%) | Seroprevalence<sup>2</sup><br>n (%; 95% CI<sup>3</sup>)(%; 95% CI) | Number sampled<br>N (%) | Seroprevalence<sup>2</sup><br>n (%; 95% CI<sup>3</sup>) |
| **All participants†** | 7010 | 5124 (73.1; 72.0-74.1) | 7510 | 6823 (90.9; 90.2-91.5) |
| **Sex** |  |  |  |  |
| Male | 2941 (42%) | 1999 (68; 66.3-69.6) | 3096 (41.4%) | 2726 (88; 86.9-89.1) |
| Female | 4065 (58%) | 3123 (76.8; 75.5-78.1) | 4390 (58.6%) | 4075 (92.8; 92.0-93.6) |
| **Age group – yr<sup>‡</sup>** |  |  |  |  |
| <12 | 753 (10.7%) | 423 (56.2; 52.6-59.7) | 584 (7.8%) | 491 (84.1; 80.9-86.8) |
| 12–18 | 622 (8.9%) | 459 (73.8; 70.2-77.1) | 553 (7.4%) | 523 (94.6; 92.4-96.2) |
| >18 to 50 | 4047 (57.7%) | 2978 (73.6; 72.2-74.9) | 4614 (61.6%) | 4204 (91.1; 90.3-91.9) |
| >50 | 1588 (22.7%) | 1264 (79.6; 77.5-81.5) | 1739 (23.2%) | 1587 (91.3; 89.8-92.5) |
| **Vaccination status<sup>‡</sup>** |  |  |  |  |
| Not vaccinated (all age groups) | 4938 (70.4%) | 3473 (70.3; 69.0-71.6) | 4891 (65.5%) | 4377 (89.5; 88.6-90.3) |
| Vaccinated | 1319 (18.8%) | 1228 (93.1; 91.6-94.3) | 1995 (26.7%) | 1918 (96.1; 95.2-96.9) |
| <12yrs | 753 (10.7%) | 423 (56.2; 52.6-59.7) | 584 (7.8%) | 491 (84.1; 80.9-86.8) |
| **Vaccination by age group** |  |  |  |  |
| <12 unvaccinated | 753 (10.7%) | 423 (56.2; 52.6-59.7) | 584 (7.8%) | 491 (84.1; 80.9-86.8) |
| 12–18 unvaccinated | 603 (8.6%) | 443 (73.5; 69.8-76.8) | 442 (5.9%) | 412 (93.2; 90.5-95.2) |
| 12–18 vaccinated | 19 (0.3%) | 16 (84.2; 62.4-94.5) | 106 (1.4%) | 106 (100; 96.5-100.0) |
| >18 to 50 unvaccinated | 3356 (47.9%) | 2335 (69.6; 68.0-71.1) | 3470 (46.5%) | 3109 (89.6; 88.5-90.6) |
| >18 to 50 vaccinated | 691 (9.9%) | 643 (93.1; 90.9-94.7) | 1130 (15.1%) | 1082 (95.8; 94.4-96.8) |
| >50 unvaccinated | 979 (14%) | 695 (71; 68.1-73.7) | 979 (13.1%) | 856 (87.4; 85.2-89.4) |
| >50 vaccinated | 609 (8.7%) | 569 (93.4; 91.2-95.1) | 759 (10.2%) | 730 (96.2; 94.6-97.3) |
| **Reported previous covid -19 positive test** |  |  |  |  |
| Never tested | 5956 (85%) | 4272 (71.7; 70.6-72.9) | 7209 (96.4%) | 6547 (90.8; 90.1-91.5) |
| Tested positive | 195 (2.8%) | 172 (88.2; 82.9-92.0) | 43 (0.6%) | 43 (100; 91.8-100.0) |
| Tested negative | 859 (12.3%) | 680 (79.2; 76.3-81.7) | 229 (3.1%) | 207 (90.4; 85.9-93.6) |
| **Smoking status<sup>¶</sup>** |  |  |  |  |
| Non-smoker | 4086 (58.3%) | 3172 (77.6; 76.3-78.9) | 4336 (58%) | 3986 (91.9; 91.1-92.7) |
| Daily | 1115 (15.9%) | 741 (66.5; 63.6-69.2) | 1331 (17.8%) | 1173 (88.1; 86.3-89.8) |
| Once or twice a week | 238 (3.4%) | 178 (74.8; 68.9-79.9) | 393 (5.3%) | 361 (91.9; 88.7-94.2) |
| Occasionally | 196 (2.8%) | 151 (77; 70.7-82.4) | 278 (3.7%) | 257 (92.4; 88.7-95.0) |
| **<18yrs** | 1375 (19.6%) | 882 (64.1; 61.6-66.6) | 1137 (15.2%) | 1014 (89.2; 87.2-90.9) |
| **Comorbidities** |  |  |  |  |
| **None** | 4631 (66.1%) | 3432 (74.1; 72.8-75.4) | 5233 (70%) | 4758 (90.9; 90.1-91.7) |
| **1 or more** | 1004 (14.3%) | 810 (80.7; 78.1-83.0) | 1111 (14.9%) | 1025 (92.3; 90.5-93.7) |
| **<18yrs (not assessed)** | 1375 (19.6%) | 882 (64.1; 61.6-66.6) | 1137 (15.2%) | 1014 (89.2; 87.2-90.9) |
| **HIV status** |  |  |  |  |
| **HIV negative** | 6460 (92.2%) | 4728 (73.2; 72.1-74.3) | 6850 (91.6%) | 6232 (91; 90.3-91.6) |
| **HIV positive** | 550 (7.8%) | 396 (72; 68.1-75.6) | 631 (8.4%) | 565 (89.5; 86.9-91.7) |

Note: Missing in current sero-survey 3: sex= 24; age-group =20; vaccination status =40; ever tested covid = 29, smoke=35; co-morbidities=29 and self-reported HIV=29.
Missing in previous sero-survey 2: sex= 4
<sup>1</sup> *Madhi et al, 2022*<sup>9</sup>
<sup>2</sup> Seroprevalence was defined as seropositive for anti-S or anti-N IgG, irrespective of vaccinations status.
3CI, confidence interval; Confidence intervals have not been adjusted for multiplicity and should not be used for inference.

Only 29.0% (n=1995/6886) of individuals older than 12 years of age who were eligible to receive a Covid-19 vaccine, received at least a single dose. Sero-positivity was slightly higher in individuals older than 12 years who had received at least a single dose of Covid-19 vaccine (96.1%; 95%CI: 95.2-96.9) compared with unvaccinated individuals (89.5%; 95%CI: 88.6-90.3). Higher sero-positivity was evident for the vaccinated compared with the unvaccinated across all age groups > 12 years, Table 2, and across districts and sub-districts; Supplementary Tables S5 and S6.

### Sero-conversion and sero-response

Restricting analyses to individuals with paired samples and no Covid-19 vaccination following pre-BA.1 sero-survey who were anti-N and anti-S IgG seronegative at the previous survey, 74.9% (95%CI: 71.0-78.5; range 61.5% to 80.5% across the districts) demonstrated sero-conversion; Table 1. High rates of sero-conversion were also observed across all stratified age-groups, ranging from 66.4% (95%CI: 57.9-74.0) in the >50-year age-group to 90.9% (95%CI: 76.4-96.9) in the 12–17-year age-group; Table 1 and Supplementary Table S7.

Based on the composite of sero-conversion or sero-response during the BA.1 dominant wave, 63.9% (95%CI; 61.8-65.9) had serological evidence of SARS-CoV-2 infection, varying from 54.1% to 68.5 across districts. The percentage with serological evidence of SARS-CoV-2 infection ranged from 59.0% (95%CI: 54.9-62.9) in the >50-year age-group to 77.5% (95%CI: 70.4-83.3) in the 12-to-17-year age-group; Table 1. Serological evidence of SARS-CoV-2 infection in the BA.1-dominant wave was higher in individuals not vaccinated against Covid-19 (67.0%; 95%CI: 64.6%-69.3%) than for individuals vaccinated in the only prior to the preceding sero-survey (54.8%; 95%CI: 50.5-59.0%), including when stratified by age-groups eligible for vaccination; Supplementary Table S8. Similar trends were seen for individuals vaccinated in the interval period between the two surveys; Supplementary Table S9.

### Covid-19 rates, hospitalizations and deaths

By June 6^th^, 2022, the daily case and hospitalization rates in the BA.4/BA.5 dominant wave had already returned to those of inter-wave period prior to the onset of the BA.4/BA.5 dominant wave, Figure 2. Compared with the BA.1 dominant wave, the BA.4/BA.5 dominant wave contributed an even lower percentage of the total cumulative number of recorded Covid-19 events since the start of the pandemic. Whereas the BA.1 dominant wave contributed to 21.3%, 14.1% and 5.9% of the cumulative number of recorded Covid-19 cases, hospitalizations and deaths, respectively; the corresponding percentages for the BA.4/BA.5 dominant wave were 8.2%, 5.1% and 1.6%; Table 3 and Figure 2. The excess mortality estimates in the BA.4/BA.5 dominant wave was higher (n=3,678) than in the BA.1 dominant wave (n=2,734) and contributed to 5.9% and 4.4% of all excess mortality deaths since the start of the pandemic, respectively.

**Table 3:** Cumulative reported Covid-19 cases, hospitalizations, recorded deaths, and excess mortality in Gauteng Province by Covid-19 wave.

| Outcomes | Pre-BA.1 dominant wave<br>cumulative | BA.1 dominant wave | BA4/5 sublineage<br>resurgence | Total |
| --- | --- | --- | --- | --- |
| Period of case wave | March 7, 2020 to Oct. 22, 2021 | Oct. 23, 2021 to March 21, 2022 | March 22, 2022 to June 6, 2022 | Not applicable |
| Inferred infections from sero-survey<sup>1</sup> | 8,391,304 | 10,167,996 |  | Not applicable |
| Cases – no.† | 926,138 | 279,510 | 108,108 | 1,313,756 |
| Cumulative Case rate per 100,000 population | 4,923 | 1,768 | 684 | 8,309 |
| Annualised case rate per 100,000 population | 3,020 | 5,358 | 2,443 | 3,693 |
| Proportion of total cumulative cases, % | 70.5 | 21.3 | 8.2 | 100 |
| Inferred infection: recorded case ratio (95%CI) | 9.1 (8.9-9.2). | 36.4 (35.1-37.6) |  | Not applicable |
| Period of Covid-19 hospitalisation wave | March 7, 2020 to Nov. 1, 2021 | Nov. 2, 2021 to March 23, 2022 | March 24, 2022, 2022 - June 6, 2022 | Not applicable |
| Hospitalizations– no.* | 127,439 | 22207 | 8094 | 157,813 |
| Cumulative Hospitalization rate per 100,000 population | 806 | 141 | 51 | 998 |
| Proportion of total cumulative hospitalizations, % | 80.8 | 14.1 | 5.1 | 100 |
| Inferred infection: recorded hospitalization ratio (95%CI) | 65.8 (64.9-66.7) | 457.9 (441.5-473.6) |  | Not applicable |
| Period of recorded Covid-19 deaths wave | March 31, 2020 to Nov. 3, 2021 | Nov. 4, 2021 to April 14, 2022 | April 15, 2022 to June 6 2022 | Not applicable |
| Recorded deaths in wave – no. | 28,026 | 1,798 | 475 | 30,304 |
| Cumulative Recorded death rate per 100,000 population<sup>5</sup> | 177.3 | 11.4 | 3.0 | 191.7 |
| Proportion of total cumulative recorded deaths, % | 92.5 | 5.9 | 1.6 | 100 |
| Inferred infection: recorded death ratio (95%CI) | 299.4 (294.9-303.5) | 5655.2 (5452.6-5849.3) |  | Not applicable |
| Infection Fatality Ratio (IFR) for recorded deaths | 0.33%. | 0.02% |  | Not applicable |
| Period of excess deaths wave | March 3, 2020 to Nov. 27, 30, 2021 | Nov. 28, 2021 to March 19, 2022 | March 20, 2022 to June 6 2022 |  |
| Excess deaths in wave – no. | 56,149 | 2,734 | 3,678 | 62,561 |
| **Cumulative Excess death rate per 100,000 population** | 355.1 | 17.3 | 22.6 | 406.7 |
| **Proportion of total cumulative excess deaths, %** | 89.8 | 4.4 | 5.9 | 100 |
| **Inferred infection: excess deaths ratio (95%CI)** | 149.4 (147.2-151.5) | 3719.1 (3585.9-3846.8) |  | Not applicable |
| **Infection Fatality Ratio<sup>2</sup> (IFR) for excess deaths** | 0.67% | 0.03% |  | Not applicable |

<sup>1</sup>The inferred number of infections in the population pre-Omicron BA.1 dominant wave was derived by multiplying the seroprevalence in unvaccinated individuals at the time of the pre- BA.1 sero-survey by the STATS-SA population<sup>13</sup>. For post- BA.1 inferred number of infections was obtained by multiplying the proportion of unvaccinated individuals showing overall serological evidence of SARS-CoV-2 infection (Table S8) between the pre- BA.1 and post-BA.1 dominant wave sero-surveys, by the STATS-SA population.
<sup>2</sup> The Infection Fatality Ratio was calculated as the inverse of the Inferred Infection to recorded deaths or excess ratios.
\*All data are from the National Institute for Communicable Diseases daily databases<sup>11</sup> except for weekly excess deaths. Excess mortality from natural causes was defined per and sourced from the South African Medical Research Council<sup>12</sup>; the excess mortality data are reported through to 4 June 2022. Other waves are lagged with respect to cases. Consequently, each of the hospitalization, recorded death, and excess death waves have their own cut-points determining the start and end of the four epidemic waves.
†Changes in testing rates, particularly the lower rates during Wave 1 due to constraints in laboratory capacity and prioritization of testing for hospitalized individuals, prevent direct comparisons, especially in terms of case numbers during the first wave in relation to the subsequent waves. Cases include asymptomatic and symptomatic individuals. Cumulative reported cases were sourced from the National Department of Health.
‡Hospitalization data are from DATCOV, hosted by the National Institute for Communicable Disease,<sup>11</sup> as described previously.<sup>9,10</sup> The system was developed during the course of the first wave, with gradual onboarding of facilities; hence, these data could underestimate hospitalized cases in the first wave relative to subsequent waves. The hospitalized cases include individuals with Covid-19, as well as coincidental infections identified as part of routine testing for SARS-CoV-2 of individuals admitted to the facilities to assist in triaging of patients in the hospital. §Cumulative reported deaths were sourced from the National Department of Health.

**Figure 2:**
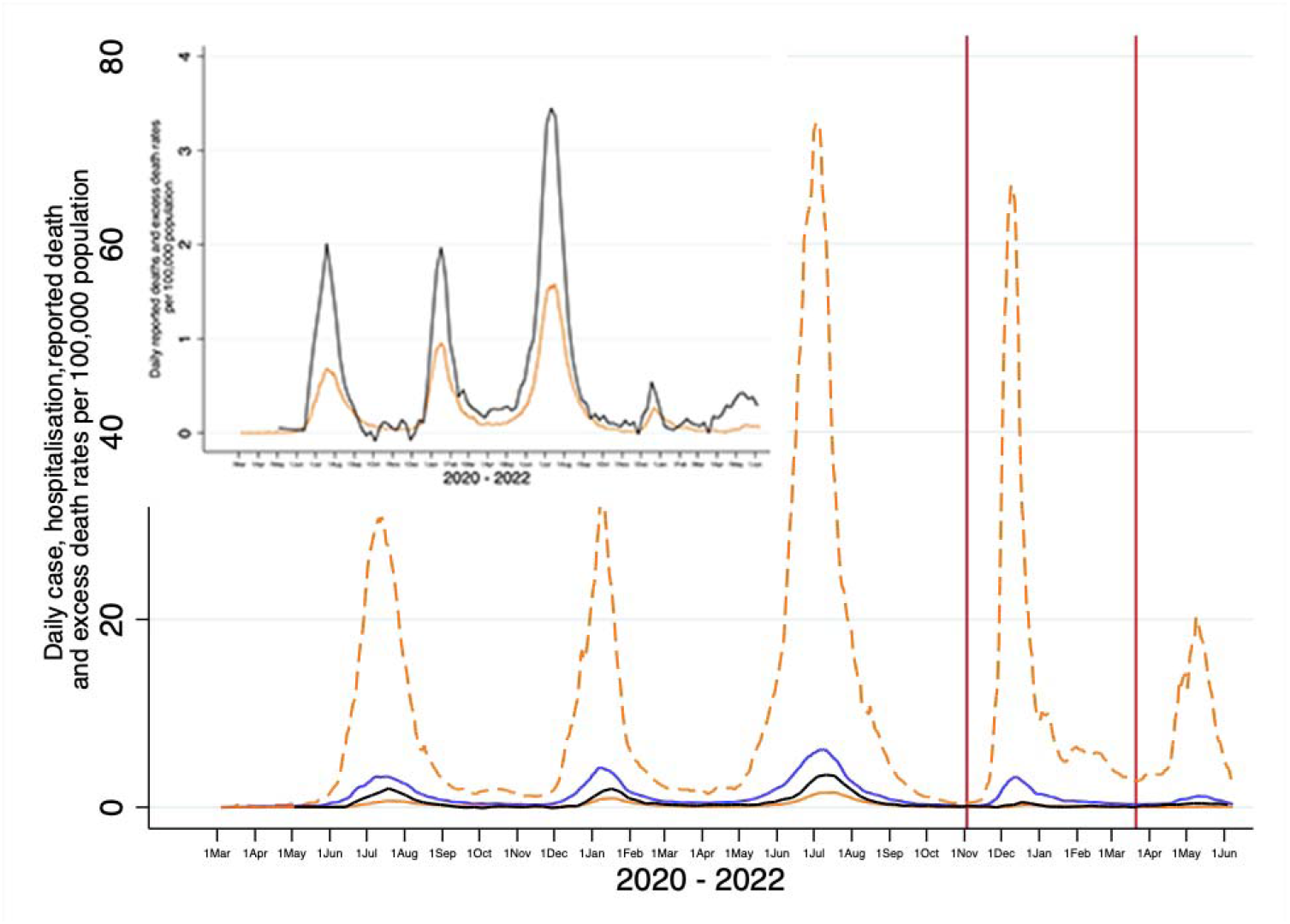
Overall trends of daily incidence per 100.000 of recorded Covid-19 cases, hospitalizations and deaths; and excess mortality attributable Covid-19 deaths for Gauteng, South Africa. Inset figure magnifies Covid-19 recorded deaths and excess mortality. For Covid-19 cases the waves periods for pre-BA.1 period cumulative, Omicron BA.1 dominant wave and BA4/5 resurgence were March 7, 2020 to Oct. 22, 2021; Oct. 23,2021 to March 21, 2022 and March 22, 2022 to June 6, 2022, respectively. For Covid-19 hospitalizations the wave periods for pre-BA.1 cumulative, Omicron BA.1 dominant wave and BA4/5 resurgence were March 7, 2020 to Nov. 1, 2021; Nov. 2, 2021 to March 23,2022 and March 24, 2022 to June 6, 2022, respectively. For Covid-19 recorded deaths the wave periods for pre-BA.1 cumulative, Omicron BA.1 dominant wave and BA4/5 resurgence were March 31, 2020 to Nov. 3, 2021; Nov. 4, 2021 to April 14, 2022 and April 14, 2022 to June 6, 2022, respectively. For Covid-19 attributable excess deaths the wave periods for pre-BA.1 cumulative period, Omicron BA.1 dominant wave and BA4/5 resurgence were March 3, 2020 to Nov. 27, 2021; Nov. 28, 2021 to March 19, 2022 and March 20, 2022 to June 6, 2022, respectively. All rates are smoothed using a 7-day moving average except for excess mortality.

The cumulative incidence rate (per 100,000) of recorded Covid-19 cases declined from 4,923 over the first three Covid-19 waves (pre-BA.1), to 1,768 in the BA.1 dominant wave, and 684 in the BA.4/BA.5 dominant wave. The inferred infections to recorded Covid-19 cases ratio increased from 9.1 in the pre-BA.1 period to 36.4 in the BA.1 dominant wave, indicating greater under-ascertainment of infections in the latter wave; Table 3.

The cumulative incidence rate (per 100,000) of Covid-19 hospitalizations declined from 806 in the pre-BA.1 period, to 141 and 51 in the BA.1 and BA.4/BA.5 dominant waves, respectively. The BA.4/BA.5 wave contributed to 5.1 % of all Covid-19 hospitalizations since the start of the pandemic, compared with 14.1% having transpired during the BA.1 dominant wave. Whereas an estimated 66 infections resulted in one Covid-19 hospitalization during the pre-BA.1 period, there was one Covid-19 hospitalization for every 458 inferred infections during the BA.1 dominant wave; Table 3.

The cumulative incidence rate (per 100,000) of recorded Covid-19 deaths declined from 177.3 to 11.4 and 3.0 during the pre-BA.1, BA.1 and BA.4/BA.5 dominant waves, respectively. Ninety-three percent of all recorded deaths since the start of the pandemic preceded the onset of the BA.1 dominant wave, 5.9% occurred during the BA.1 dominant wave and only 1.6% in the BA.4/BA.5 dominate wave. Overall, there was one recorded Covid-19 death for every 299 and 5655 inferred infections, with IFR of 0.33% and 0.02% in the pre-BA.1 period and BA.1 dominant wave, respectively; Table 3.

The cumulative incidence rate (per 100,000) of Covid-19 attributable deaths using the excess mortality estimates was approximately 2.1-fold higher compared with the recorded deaths (406.7 vs. 191.7, respectively). The cumulative incidence rate of Covid-19 attributable (excess) deaths declined from 355.1 over the pre-BA.1 period to 17.3 over the BA.1 dominant wave and is 22.6 in the BA.4/BA.5 dominant wave. Overall, there was one death for every 149.4 and 3719.1 inferred infections corresponding to IFRs of 0.67% and 0.03% in the pre-BA.1 period and BA.1 dominant waves, respectively; Table 3.

Age-group stratified analysis of the cumulative incidence rates showed the same downward trend of recorded Covid-19 cases, hospitalizations and deaths from the pre-BA.1 period, during the BA.1 and BA.4/BA.5 dominant waves, Figures 3 a-d and Supplementary Table S10. In adults older than 50 years of age, the Omicron BA.1 and BA.4/BA.5 dominant waves respectively contributed to 9.5% and 3.5% of Covid-19 hospitalizations and 5.3% and 0.8% of recorded deaths which occurred since the start of the pandemic. Also comparing the pre-BA.1 period with the BA.1 dominant wave, the ratio of inferred infections for every Covid-19 hospitalisation increased from 13 to 209; and from 38 to 1289 for recorded Covid-19 deaths corresponding to IFR of 2.66% and 0.08%, Supplementary Table S10.

**Figure 3 a-d:**
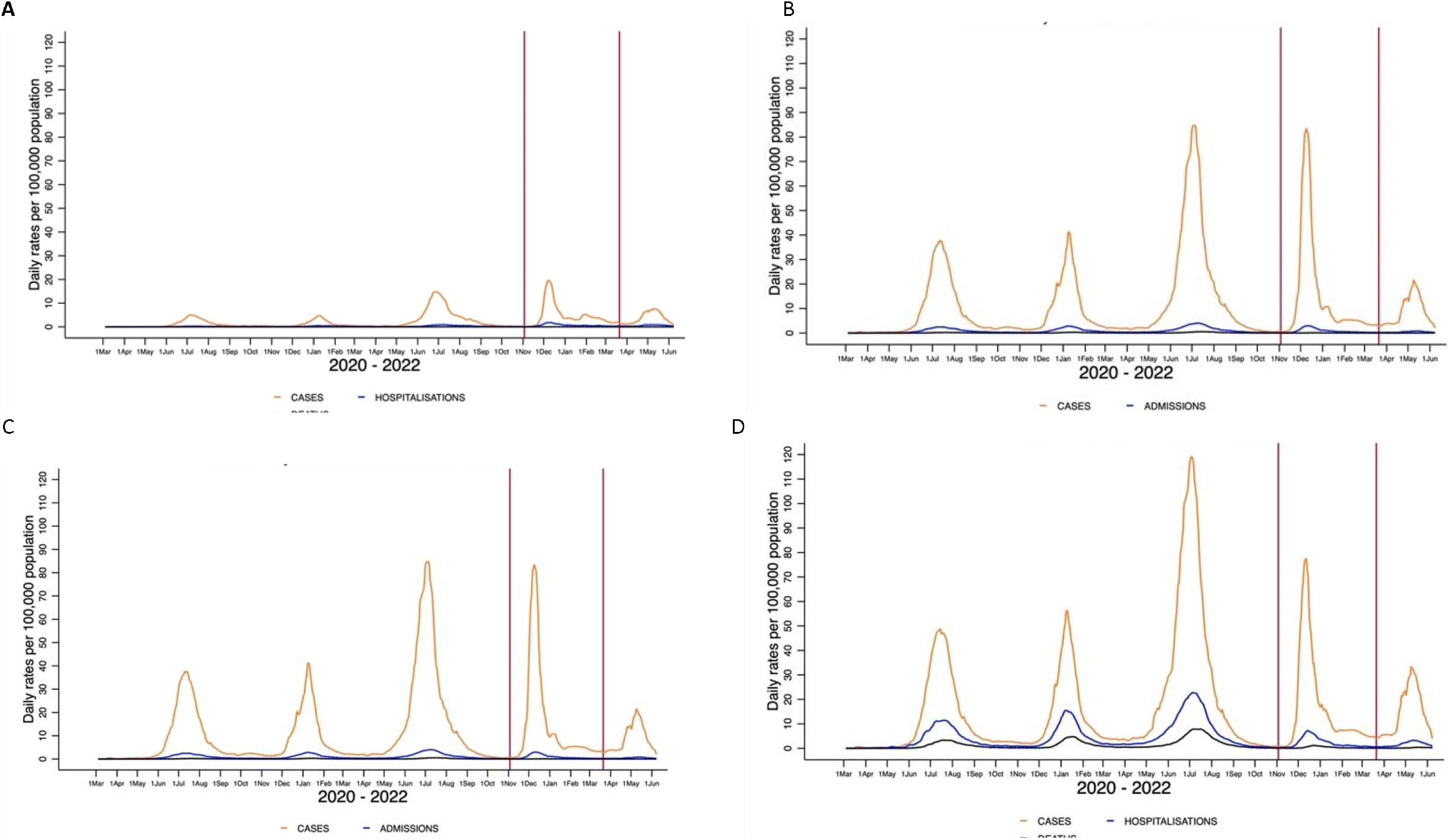
Agegroup stratified analysis of daily moving averages of recorded cases, hospitalizations and deaths in Gauteng Province for each of five Covid-19 waves. For Covid-19 cases the waves periods for pre-Omicron BA.1 cumulative, Omicron BA.1 dominant wave and BA4/5 resurgence were March 7, 2020 to Oct. 22, 2021; Oct. 23,2021 to March 21, 2022 and March 22, 2022 to June 6, 2022, respectively. For Covid-19 hospitalizations the wave periods for pre-BA.1 cumulative, Omicron BA.1 dominant wave and BA4/5 resurgence were March 7, 2020 to Nov. 1, 2021; Nov. 2, 2021 to March 23,2022 and March 24, 2022, 2022 - June 6, 2022, respectively. For Covid-19 recorded deaths the wave periods for pre-BA.1 cumulative, Omicron BA.1 dominant wave and BA4/5 resurgence were March 31, 2020 to Nov. 3, 2021; Nov. 4, 2021 to April 14, 2022 and April 15, 2022 to June 6 2022, respectively. All rates are smoothed using a 7-day moving average.

## DISCUSSION

Despite only 26.7% (1995/7470) of individuals in the survey having received at least a single dose of Covid-19 vaccine at the time of sampling, the overall sero-positivity for SARS-CoV-2 was 90.9% after the BA.1 dominant wave had subsided in Gauteng; including 89.5% in unvaccinated individuals older than 12 years of age. Using paired serology data, 63.9% of the population were infected with the BA.1 variant during the fourth Covid-19 wave in Gauteng. Serological evidence of infection during the BA.1 dominant wave was higher (67.0%) in individuals Covid-19 unaccinated compared with those who had only been vaccinated before the onset of the BA.1 doinant wave (54.8%). Against this background of high sero-positivity and high rates of BA.1 infections, we observed further decoupling of SARS-CoV-2 infection and recorded Covid-19 hospitalizations and deaths during the subsequent BA.4/BA.5 dominant wave which has now subsided in Gauteng. Notably, the BA.4/BA.5 dominant wave contributed to 5.1% and 1.6% of all recorded Covid-19 hospitalizations and deaths since the start of the pandemic, compared with 14.1% and 5.9% of these events during the BA.1 dominant wave.

In the current analysis, we are also able to provide an update on the burden of Covid-19 during the BA.1 dominant wave compared with the earlier three Covid-19 waves. The SARS-CoV-2 cumulative IFR declined from 0.33% to 0.02% for recorded deaths, and from 0.67% to 0.03% for attributable Covid-19 deaths based on excess mortality estimates in the pre-BA.1 waves compared with the BA.1 dominant wave. In comparison, the IFR for seasonal influenza virus pre Covid-19 pandemic in South Africa is estimated to be 0.05%, based conservatively on approximately 35% (n=20.9 million) of the population with serological evidence of infection and excess mortality attributable influenza deaths of 11,000 per annum^14-16^. We may, however, have over-estimated the IFR in the pre-BA.1 period, as the cross sectional sero-prevalence used to infer infections over the course of the first three Covid-19 waves would have missed re-infections.

Notably, although there has been a 3.8-fold decrease in recorded Covid-19 deaths between the BA.1 (n=1798) and BA.4/BA.5 dominant waves (n=475); the excess mortality deaths were higher during the BA.4/BA.5 wave (3678 vs 2734). The fold difference of excess deaths to recorded Covid-19 deaths increased from 1.52 (2734/1798) in the BA.1 dominant wave to 7.74 (3678/475) in the BA.4/BA.5 dominant wave. It is unlikely there has been differential under-reporting of Covid-19 deaths between these two periods to explain this discrepancy, as there were no changes in access to healthcare and DATCOV surveillance has been stable during this time. Rather, these data suggest that excess mortality calculations are becoming less reliable in identifying Covid-19 attributable deaths. This could be due to a number of factors, including re-emergence of other infectious diseases. The BA.4/BA.5 dominant wave in Gauteng coincided with an epidemic of respiratory syncytial virus, as well as an influenza virus epidemic which occurred earlier than anticipated based on pre-Covid-19 seasonal epidemiology^17,18^. By contrast, seasonal influenza was largely absent during 2020 and there was only a mild influenza season in 2021 in South Africa. Other chronic causes of death aggravated by reduced effectiveness of the health care system during the course of the Covid-19 pandemic could also have contributed to variation over time in the consistency excess mortality estimates as a proxy for Covid-19 attributable deaths. Other limitations of our study are elaborated upon in the supplementary appendix.

In conclusion, there is uncertainty as to whether the Omicron VOC is intrinsically less virulent than earlier variants^19,20^. The propensity of BA.1 to infect the upper rather than the lower airways could also have contributed to decoupling of infection and Covid-19.^21^ Nevertheless, our study indicates that unless future variants harbour mutations which evade poly-epitopic CD4^+^ and CD8^+^ immunity induced by current vaccines and past infection, or the virus becomes intrinsically more virulent, Covid-19 no longer poses a major threat of a large burden of severe disease and death compared with the period before the evolution of extensive population immunity. Notably, the breadth of T-cell immunity against SARS-CoV-2 is expected to be more diverse in settings such as South Africa where there has been a high force of SARS-CoV-2 infection^22^. Poly-epitopic T-cell responses following SARS-CoV-2 infections by all variants to date are directed against the spike, nucleocapsid and membrane protein epitopes. These responses contribute to attenuating the progression of infection to severe disease and may also reduce transmission of the virus^6^. Although hybrid immunity with three doses of Covid-19 vaccines protects better against BA.1 symptomatic Covid-19 than infection-only immunity, both are similarly effective against severe Covid-19^17^. The very high rates of infection and re-infection amongst both the unvaccinated and vaccinated decoupled from severe disease in a population with very high sero-positivity prevalence, herald an endemic phase to the pandemic akin to other endemic respiratory viruses which may cause epidemics such as seasonal influenza. Given a likely future scenario of continued infection and re-infection in all populations globally, there is, however, the possibility of an emergent more virulent and immune-evasive variant, necessitating ongoing surveillance.

## Data Availability

All data produced in the present work are contained in the manuscript

## ACKNOWLEDGEMENTS

We thank the epidemiology team (Andronica Moipone Shonhiwa, Genevie Ntshoe, Joy Ebonwu, Lactatia Motsuku, Liliwe Shuping, Mazvita Muchengeti, Jackie Kleynhans, Gillian Hunt, Victor Odhiambo Olago, Husna Ismail, Nevashan Govender, Ann Mathews, Vivien Essel, Veerle Msimang, Tendesayi Kufa-Chakezha, Nkengafac Villyen Motaze, Natalie Mayet, Tebogo Mmaborwa Matjokotja, Mzimasi Neti, Tracy Arendse, Teresa Lamola, Itumeleng Matiea, Darren Muganhiri, Babongile Ndlovu, Khuliso Ravhuhali, Emelda Ramutshila, Salaminah Mhlanga, Akhona Mzoneli, Nimesh Naran, Trisha Whitbread, Mpho Moeti, Chidozie Iwu, Eva Mathatha, Fhatuwani Gavhi, Masingita Makamu, Matimba Makhubele, Simbulele Mdleleni, Bracha Chiger, Jackie Kleynhans, and Michelle Groome) and the information technology team (Tsumbedzo Mukange, Trevor Bell, Lincoln Darwin, Fazil McKenna, Ndivhuwo Munava, Muzammil Raza Bano, and Themba Ngobeni) at the National Institute for Communicable Diseases Notifiable Medical Conditions Surveillance System. The authors would like to acknowledge Gauteng Department of Health for supporting field operations.

## DATA SHARING

Data are available at www.wits-vida.org; requests for data sharing should be directed to Professor Shabir A. Madhi

