## Supplemental material for "SARS-CoV-2 infections during Omicron (BA.1) dominant wave and subsequent population immunity in Gauteng, South Africa"

**Supplementary Appendix**

| **Contents** | **Page** |
| --- | --- |
| **Supplementary Methods** | **3** |
| Study setting and sero-survey | 3 |
| Serology assays | 3 |
| Study limitations | 3 |
| **Supplementary Tables** |  |
| Supplementary Table S1: Time period over which sero-survey was undertaken in different sub-districts in Gauteng, South Africa. | 5 |
| Supplementary Table S2a: Vaccination classification of individuals with paired samples for immunoglobulin G analysis at the pre-BA.1 and post-BA.1 sero-surveys. | 6 |
| Supplementary Table S2b: Criteria used to determine seroconversion or seroresponse in the pre-Omicron and post Omicron BA.1 sero-survey interval period | 6 |
| Supplementary Table S3: Demographic characteristics and anti-nucleocapsid or anti-spike protein IgG seroprevalence in individuals sampled in only post-BA.1 sero-survey and those samples in both pre-BA.1 and post-BA.1 sero-surveys. | 7 |
| Supplementary Table S4: Seroprevalence of SARS-CoV-2 anti-spike (anti-S) or anti-nucleocapsid (anti-N) immunoglobulin G (IgG) Gauteng Province across the Districts and sub-Districts irrespective of Covid-19 vaccination status. | 9 |
| Supplementary Table S5: Seroprevalence of SARS-CoV-2 anti-spike (anti-S) or anti-nucleocapsid (anti-N) immunoglobulin G (IgG) Gauteng Province across the Districts and sub-Districts vaccinated individuals older than 12 years only. | 10 |
| Suppl Table S6: Seroprevalence of SARS-CoV-2 anti-spike (anti-S) or anti-nucleocapsid (anti-N) immunoglobulin G (IgG) Gauteng Province across the Districts and sub-Districts unvaccinated individuals older than 12 years only. | 11 |
| Supplementary Table S7: Sero-response to SARS-CoV-2 anti-spike (anti-S) and anti-nucleocapsid (anti-N) immunoglobulin G (IgG) in paired samples from individuals who did not receive any Covid-19 vaccine following the pre-BA.1 sero-survey stratified by district, age-groups and vaccination status at pre-BA.1 sero-survey. | 12 |
| Supplementary Table S8: Seroconversion, sero-response and overall serological evidence of SARS-CoV-2 infection between the pre-BA.1 and post-BA.1 sero-surveys in individuals who did not receive any Covid-19 vaccine following the pre-BA.1 sero-survey and stratified by vaccination status at pre-Omicron sero-survey. | 13 |
| Supplementary Table S9: Seroconversion, sero-response and overall serological evidence of SARS-CoV-2 infection between the pre-BA.1 and post-BA.1 sero-surveys in individuals who received any Covid-19 vaccine following the pre-BA.1 sero-survey and stratified by vaccination status at pre-BA.1 sero-survey. | 15 |
| Supplementary Table S10: Cumulative reported Covid-19 cases, hospitalizations, recorded deaths, and excess mortality in Gauteng Province by age-group and Covid-19 wave | 16 |
| **Supplementary Figures** |  |
| Supplementary Figure S1: Seroprevalence in pre- and post-Omicron serosurvey in unvaccinated individuals | 19 |
| **Supplementary References** | 20 |

**SUPPLEMENTARY METHODS**

**Study setting and sero-survey**

Gauteng is home to 26% (n=15.5 million) of South Africa’s population. The sero-survey included households involved in the two earlier surveys. Nevertheless, based on availability and willingness to participate, the study design allowed for replacement households and different individuals within the same household which was previously involved to participate in the separate surveys. Details of the sampling framework, data collection tools and sample size calculation to enable adequate power to evaluate sero-positivity for SARS-CoV-2 at the district and sub-district levels has been previously described^1^. As done with the preceding survey, there was a 10% increase in the households that were sampled in each of the pre-defined clusters to account for possible nonparticipation, out-migration, and death since the previous survey.

**Serology assays**

Binding IgG antibodies were measured by bead-based immunoassays on the Luminex platform to quantify serum IgG binding to full length spike and nucleocapsid; and reported in binding antibody units per millilitre (BAU/ml). The anti-N and anti-S assay was calibrated against a research reagent (NIBSC 20/130) and first WHO International Standard for anti-SARS-CoV-2 (NIBSC 20/136) for anti-SARS-CoV-2 antibody obtained from the National Institute for Biological Standards and Control, UK).

**DISCUSSION**

**Study limitations:**

Limitations of our study include that we were unable to adequately infer the number of SARS-CoV-2 infections during the first three Covid-19 waves, as the use of a single cross-sectional survey pre-BA.1 dominant would have not accounted for re-infections which may have transpired. Although we did an earlier survey at the time of the peak of the second wave due to the Beta VOC, we used a different anti-RBD assay which was subsequently shown not to be as sensitive as the composite of the anti-N and anti-S assays in detecting previous infections. Hence, we may have over-estimated the infection to case ratios in the pre-BA.1 dominant era, as well as possibly the IFR. It is possible that individuals who were anti-N and anti-IgG negative at the pre-BA.1 survey may have been previously infected but were sero-negative due to waning of IgG to un-detectable levels, hence the group should not be regarded as being SARS-CoV-2 naïve at any stage in our survey. Also, our study is unable to adjust for different interventions which have undergone continuous but variable changes during the course of the pandemic, which could have affected susceptibility to infections, as well as progression to severe disease. Nevertheless, South Africa response to the BA.1 dominant wave was to remain at the lowest level of restrictions of activity in society, which has remained the case even with the BA.4/BA.5 dominant wave. Also, access to health care, including hospitalization has been less of an issue during the course of the BA.1 and BA.4/BA.5 dominant wave compared with the earlier waves when health care facilities were overwhelmed which could have resulted in higher case fatality risks in earlier waves.

Another limitation of the analysis is that we used serological evidence of SARS-CoV-2 infection in the Covid-19 unvaccinated population (67.0%) to infer the number of cases of infection which transpired during the BA.1 dominant wave during the interval period between the pre-BA.1 and after the BA.1 dominant wave. This was done to minimise any confounding which receipt of Covid-19 could have had on using kinetics of antibody changes to infer serological evidence of infections. Nevertheless, at the overall population level, serological evidence of infection was 63.9%, hence, our approach is unlikely to have significantly increased the estimated number of inferred cases.

Another limitation is that the extreme infectiousness of Omicron and its sub-lineages could have resulted in significant misclassification of Covid-19 hospitalisation and reported death data, as the DATACov surveillance as it has not been possible to distinguish between those admitted or dying with SARS-CoV-2 infection, as opposed to dying from Covid-19. Consequently, the recorded Covid-19 hospitalizations and deaths during the BA.1 and BA.4/BA.5 dominant waves may be over-estimating the burden of severe Covid-19 during these waves compared with pre-BA.1 period.

**Supplementary Table S1: Time period over which sero-survey was undertaken in different sub-districts in Gauteng, South Africa.**

| Sub-district | survey timelines | |
| --- | --- | --- |
| Johannesburg A | 01-Mar-22 | 24-Mar-22 |
| Johannesburg B | 24-Mar-22 | 11-Apr-22 |
| Johannesburg C | 01-Mar-22 | 04-Apr-22 |
| Johannesburg D | 01-Mar-22 | 30-Mar-22 |
| Johannesburg E | 01-Apr-22 | 07-Apr-22 |
| Johannesburg F | 01-Mar-22 | 06-Apr-22 |
| Johannesburg G | 02-Mar-22 | 07-Apr-22 |
| Ekurhuleni E1 | 02-Mar-22 | 05-Apr-22 |
| Ekurhuleni E2 | 02-Mar-22 | 25-Mar-22 |
| Ekurhuleni N1 | 14-Mar-22 | 06-Apr-22 |
| Ekurhuleni N2 | 01-Mar-22 | 08-Apr-22 |
| Ekurhuleni S1 | 14-Mar-22 | 24-Mar-22 |
| Ekurhuleni S2 | 01-Mar-22 | 28-Mar-22 |
| Emfuleni | 02-Mar-22 | 26-Mar-22 |
| Lesedi | 31-Mar-22 | 07-Apr-22 |
| Midvaal | 28-Mar-22 | 01-Apr-22 |
| Tshwane Region 1 | 02-Mar-22 | 07-Apr-22 |
| Tshwane Region 2 | 15-Mar-22 | 01-Apr-22 |
| Tshwane Region 3 | 31-Mar-22 | 07-Apr-22 |
| Tshwane Region 4 | 01-Apr-22 | 04-Apr-22 |
| Tshwane Region 5 | 02-Mar-22 | 21-Mar-22 |
| Tshwane Region 6 | 20-Mar-22 | 06-Apr-22 |
| Tshwane Region 7 | 30-Mar-22 | 01-Apr-22 |
| Merafong City | 01-Mar-22 | 13-Mar-22 |
| Mogale City | 20-Mar-22 | 02-Apr-22 |
| Rand West City | 01-Mar-22 | 07-Apr-22 |

**Supplementary Table S2a: Vaccination classification of individuals with paired samples for immunoglobulin G analysis at the pre-BA.1 and post-BA.1 sero-surveys.**

| Covid-19 vaccination history reported in pre-Omicron and post Omicron BA.1^1^ | Count | Covid-19 vaccine classification | |
| --- | --- | --- | --- |
|  |  | Vaccinated prior to pre-BA.1 survey | Vaccinated between  pre-BA.1 and post-BA.1 survey |
| Not vaccinated | 1316 | No | No |
| Not vaccinated at the pre-Omicron survey and vaccination unknown at post Omicron BA.1 survey | 222 |  |  |
| Not vaccinated at the pre-Omicron survey, vaccinated at post Omicron BA.1 survey and vaccination date prior to pre-Omicron survey | 54 | Yes |  |
| Vaccinated at the pre-Omicron survey and not vaccinated at post Omicron BA.1 survey | 180 |  |  |
| Vaccinated at the pre-Omicron survey and vaccinated at post Omicron BA.1 survey with dates unknown | 84 |  |  |
| Vaccinated at the pre- Omicron survey and vaccination at post Omicron BA.1 survey unknown | 3 |  |  |
| Vaccinated at the pre-Omicron survey, vaccinated at post Omicron BA.1 survey and vaccination date prior to pre-Omicron survey | 199 |  |  |
| Not vaccinated at the pre-Omicron survey and vaccinated at post Omicron BA.1 survey with dates unknown | 157 | No | Yes |
| Not vaccinated at the pre-Omicron survey and vaccinated between pre-Omicron and post Omicron BA.1 survey | 115 |  |  |
| Vaccinated at the pre-Omicron survey and vaccinated between pre-Omicron and post Omicron BA.1 survey | 90 | Yes |  |

^1^Vaccination was self-reported and may contain inconsistencies (for example, participant reported vaccination pre-Omicron BA.1 survey and subsequently reported never vaccinated post-Omicron BA.1 survey).

**Supplementary Table S2b: Criteria used to determine seroconversion or seroresponse in the pre-Omicron and post Omicron BA.1 sero-survey interval period**

|  | Anti-N and anti-S serostatus at pre-Omicron survey | | Anti-N or anti-S response at post Omicron BA.1 survey | Seroconversion or sero-response classification |
| --- | --- | --- | --- | --- |
|  | Anti-N IgG | Anti-S IgG |  |  |
| Individuals not vaccinated between the pre-Omicron and post Omicron BA.1 sero-surveys | Neg | Neg | Anti-N IgG+ | Sero-conversion |
|  | Neg | Neg | Anti-S IgG+ |  |
|  | Neg | Pos | ≥2 fold increase in anti-S IgG | Sero-response |
|  | Neg | Pos | Anti-N IgG+ |  |
|  | Pos | Neg | ≥2 fold increase in anti-N IgG |  |
|  | Pos | Neg | Anti-S IgG+ |  |
|  | Pos | Pos | ≥2 fold increase in anti-N IgG |  |
|  | Pos | Pos | ≥2 fold increase in anti-S IgG |  |
| Individuals vaccinated between the pre-Omicron and post Omicron BA.1 sero-surveys | Neg | Neg | Anti-N IgG+ | Sero-conversion |
|  | Neg | Pos | Anti-N IgG+ | Sero-response |
|  | Pos | Neg | ≥2 fold increase in anti-N IgG |  |
|  | Pos | Pos | ≥2 fold increase in anti-N IgG |  |

Anti-N IgG= anti-nucleopcapsid immunoglobulin G; anti-S –anti-Spike immunoglobulin G.

**Supplementary Table S3: Demographic characteristics and anti-nucleocapsid or anti-spike protein IgG seroprevalence in individuals sampled in only post-BA.1 sero-survey and those samples in both pre-BA.1 and post-BA.1 sero-surveys.**

| Category | Only sampled post-BA.1 sero-survey | | Sampled in pre-BA.1 and post-BA.1 ssero-survey | |
| --- | --- | --- | --- | --- |
|  | **Number sampled**  **N (%)** | **Seroprevalence^1^**  **n (%; 95% CI^2^)** | **Number sampled**  **N (%)** | **Seroprevalence^1^**  **n (%; 95% CI^2^)** |
| All participants† | 5090 | 4594 (90.3; 89.4-91.0) | 2420 | 2229 (92.1; 91.0-93.1) |
| Sex: Male | 2175 (42.8%) | 1900 (87.4; 85.9-88.7) | 944 (39%) | 826 (89.7; 87.6-91.5) |
| Female | 2906 (57.2%) | 2686 (92.4; 91.4-93.3) | 1474 (61%) | 1389 (93.6; 92.2-94.7) |
| Age group – yr^‡^ |  |  |  |  |
| <12 | 384 (7.6%) | 319 (83.1; 79.0-86.5) | 206 (8.5%) | 172 (86; 80.5-90.1) |
| 12–18 | 361 (7.1%) | 338 (93.6; 90.6-95.7) | 200 (8.3%) | 185 (96.4; 92.7-98.2) |
| >18 to 50 | 3300 (64.9%) | 2984 (90.4; 89.4-91.4) | 1318 (54.5%) | 1220 (92.8; 91.3-94.1) |
| >50 | 1039 (20.4%) | 948 (91.2; 89.4-92.8) | 696 (28.8%) | 639 (91.3; 89.0-93.2) |
| Vaccination status^‡^ |  |  |  |  |
| Not vaccinated (all ages) | 3395 (66.9%) | 3015 (88.8; 87.7-89.8) | 1658 (68.5%) | 1362 (91; 89.5-92.4) |
| Vaccinated | 1296 (25.5%) | 1246 (96.1; 94.9-97.1) | 556 (23%) | 672 (96.1; 94.4-97.3) |
| <12yrs | 384 (7.6%) | 319 (83.1; 79.0-86.5) | 206 (8.5%) | 172 (86; 80.5-90.1) |
| Vaccination by age group |  |  |  |  |
| <12 unvaccinated | 384 (7.6%) | 319 (83.1; 79.0-86.5) | 206 (8.5%) | 172 (86; 80.5-90.1) |
| 12–18 unvaccinated | 291 (5.7%) | 268 (92.1; 88.4-94.7) | 191 (7.9%) | 144 (95.4; 90.7-97.7) |
| 12–18 vaccinated | 70 (1.4%) | 70 (100; 94.8-100.0) | 9 (0.4%) | 36 (100; 90.4-100.0) |
| >18 to 50 unvaccinated | 2521 (49.7%) | 2239 (88.8; 87.5-90.0) | 1059 (43.8%) | 870 (91.7; 89.7-93.3) |
| >18 to 50 vaccinated | 770 (15.2%) | 736 (95.6; 93.9-96.8) | 259 (10.7%) | 346 (96.1; 93.6-97.7) |
| >50 unvaccinated | 583 (11.5%) | 508 (87.1; 84.2-89.6) | 408 (16.9%) | 348 (87.9; 84.3-90.7) |
| >50 vaccinated | 456 (9%) | 440 (96.5; 94.4-97.8) | 288 (11.9%) | 290 (95.7; 92.8-97.5) |
| Reported previous covid -19 positive test |  |  |  |  |
| Never tested | 4903 (96.6%) | 4424 (90.2; 89.4-91.0) | 2047 (84.6%) | 2123 (92.1; 90.9-93.1) |
| Tested positive | 31 (0.6%) | 31 (100; 89.0-100.0) | 69 (2.9%) | 12 (100; 75.8-100.0) |
| Tested negative | 143 (2.8%) | 127 (88.8; 82.6-93.0) | 304 (12.6%) | 80 (93; 85.6-96.8) |
| Smoking status^¶^ |  |  |  |  |
| Non-smoker | 2954 (58.2%) | 2699 (91.4; 90.3-92.3) | 1474 (60.9%) | 1287 (93.1; 91.7-94.3) |
| Daily | 920 (18.1%) | 810 (88; 85.8-90.0) | 369 (15.2%) | 363 (88.3; 84.9-91.1) |
| Once or twice a week | 271 (5.3%) | 245 (90.4; 86.3-93.4) | 107 (4.4%) | 116 (95.1; 89.7-97.7) |
| Occasionally | 185 (3.6%) | 169 (91.4; 86.4-94.6) | 64 (2.6%) | 88 (94.6; 88.0-97.7) |
| <18yrs | 745 (14.7%) | 657 (88.2; 85.7-90.3) | 406 (16.8%) | 357 (91.1; 87.8-93.5) |
| Comorbidities |  |  |  |  |
| None | 3637 (71.6%) | 3288 (90.4; 89.4-91.3) | 1571 (64.9%) | 1470 (92.1; 90.7-93.3) |
| 1 or more | 695 (13.7%) | 637 (91.7; 89.4-93.5) | 443 (18.3%) | 388 (93.3; 90.4-95.3) |
| <18yrs (not assessed) | 745 (14.7%) | 657 (88.2; 85.7-90.3) | 406 (16.8%) | 357 (91.1; 87.8-93.5) |
| HIV status |  |  |  |  |
| HIV negative | 4640 (91.4%) | 4188 (90.3; 89.4-91.1) | 2223 (91.9%) | 2044 (92.5; 91.3-93.5) |
| HIV positive | 437 (8.6%) | 394 (90.2; 87.0-92.6) | 197 (8.1%) | 171 (88.1; 82.8-92.0) |

Note: Missing in post-Omicron BA.1 sero-survey: sex= 9; age-group =6; vaccination status =15; ever tested covid = 15, smoke=15; co-morbidities=13 and self-reported HIV=13.

Missing in both pre-Omicron and post-Omicron BA.1 sero-survey: sex= 4

^1^Seroprevalence was defined as seropositive for anti-S or anti-N IgG, irrespective of vaccinations status.

^2^CI, confidence interval; Confidence intervals have not been adjusted for multiplicity and should not be used for inference.

**Supplementary Table S4: Seroprevalence of SARS-CoV-2 anti-spike (anti-S) or anti-nucleocapsid (anti-N) immunoglobulin G (IgG) Gauteng Province across the Districts and sub-Districts irrespective of Covid-19 vaccination status.**

| District / sub-district | Population total | Pre-BA.1 sero-survey | | Post-BA.1 sero-survey | |
| --- | --- | --- | --- | --- | --- |
|  |  | **Number sampled** | **Seroprevalence^1^**  **n (%; 95% CI^2^)** | **Number sampled** | **Seroprevalence^1^**  **n (%; 95% CI^2^)** |
| Anti-S or anti-Gauteng Province | **15,176,113** | 7010 | 5124 (73.1; 72.0-74.1) | 7510 | 6823 (90.9; 90.2-91.5) |
| Johannesburg District | **5,606,238** | 2468 | 1880 (76.2; 74.5-77.8) | 2630 | 2412 (91.7; 90.6-92.7) |
| Johannesburg A | 779,519 | 333 | 246 (73.9; 68.9-78.3) | 358 | 325 (90.8; 87.3-93.4) |
| Johannesburg B | 435,241 | 197 | 169 (85.8; 80.2-90.0) | 249 | 239 (96; 92.8-97.8) |
| Johannesburg C | 799,980 | 444 | 363 (81.8; 77.9-85.1) | 476 | 455 (95.6; 93.3-97.1) |
| Johannesburg D | 1,396,243 | 646 | 472 (73.1; 69.5-76.3) | 647 | 572 (88.4; 85.7-90.7) |
| Johannesburg E | 601,433 | 161 | 117 (72.7; 65.3-79.0) | 171 | 160 (93.6; 88.8-96.4) |
| Johannesburg F | 751,484 | 243 | 185 (76.1; 70.4-81.1) | 277 | 249 (89.9; 85.8-92.9) |
| Johannesburg G | 842,339 | 444 | 328 (73.9; 69.6-77.7) | 452 | 412 (91.2; 88.2-93.4) |
| Ekurhuleni District | **3,825,650** | 1861 | 1382 (74.3; 72.2-76.2) | 2132 | 1982 (93; 91.8-94.0) |
| Ekurhuleni E1 | 626 517 | 353 | 242 (68.6; 63.5-73.2) | 374 | 332 (88.8; 85.2-91.6) |
| Ekurhuleni E2 | 455 262 | 252 | 190 (75.4; 69.7-80.3) | 288 | 273 (94.8; 91.6-96.8) |
| Ekurhuleni N1 | 708 290 | 358 | 244 (68.2; 63.2-72.8) | 378 | 344 (91; 87.7-93.5) |
| Ekurhuleni N2 | 697 175 | 258 | 206 (79.8; 74.5-84.3) | 244 | 238 (97.5; 94.7-98.9) |
| Ekurhuleni S1 | 673 758 | 210 | 172 (81.9; 76.1-86.5) | 243 | 229 (94.2; 90.6-96.5) |
| Ekurhuleni S2 | 664 648 | 430 | 328 (76.3; 72.0-80.1) | 605 | 566 (93.6; 91.3-95.2) |
| Sedibeng District | **1,084,503** | 564 | 398 (70.6; 66.7-74.2) | 624 | 557 (89.3; 86.6-91.5) |
| Lesedi | 127 419 | 408 | 293 (71.8; 67.3-76.0) | 443 | 399 (90.1; 86.9-92.5) |
| Midvaal | 126 285 | 104 | 65 (62.5; 52.9-71.2) | 107 | 96 (89.7; 82.5-94.2) |
| Emfuleni | 830 798 | 52 | 40 (76.9; 63.9-86.3) | 74 | 62 (83.8; 73.8-90.5) |
| City of Tshwane District | **3,709,635** | 1464 | 975 (66.6; 64.1-69.0) | 1455 | 1255 (86.3; 84.4-87.9) |
| Tshwane 1 | 1 032 885 | 471 | 298 (63.3; 58.8-67.5) | 470 | 385 (81.9; 78.2-85.1) |
| Tshwane 2 | 436 950 | 175 | 103 (58.9; 51.5-65.9) | 175 | 144 (82.3; 76.0-87.2) |
| Tshwane 3 | 730 788 | 229 | 177 (77.3; 71.4-82.2) | 137 | 122 (89.1; 82.7-93.3) |
| Tshwane 4 | 482 448 | 78 | 46 (59; 47.9-69.2) | 97 | 83 (85.6; 77.2-91.2) |
| Tshwane 5 | 119 190 | 204 | 129 (63.2; 56.4-69.6) | 236 | 212 (89.8; 85.3-93.1) |
| Tshwane 6 | 768 446 | 245 | 175 (71.4; 65.5-76.7) | 254 | 229 (90.2; 85.9-93.2) |
| Tshwane 7 | 138 928 | 62 | 47 (75.8; 63.8-84.8) | 86 | 80 (93; 85.6-96.8) |
| West Rand District | **950,088** | 653 | 489 (74.9; 71.4-78.1) | 669 | 617 (92.2; 89.9-94.0) |
| Mogale City | 435 254 | 149 | 95 (63.8; 55.8-71.0) | 162 | 146 (90.1; 84.6-93.8) |
| Rand West City | 300 960 | 261 | 208 (79.7; 74.4-84.1) | 261 | 246 (94.3; 90.7-96.5) |
| Merafong City | 213 874 | 243 | 186 (76.5; 70.8-81.4) | 246 | 225 (91.5; 87.3-94.3) |

^1^Seroprevalence was defined as seropositive for anti-S or anti-N IgG, irrespective of vaccinations status.

^2^CI, confidence interval; Confidence intervals have not been adjusted for multiplicity and should not be used for inference.

**Supplementary Table S5: Seroprevalence of SARS-CoV-2 anti-spike (anti-S) or anti-nucleocapsid (anti-N) immunoglobulin G (IgG) Gauteng Province across the Districts and sub-Districts vaccinated individuals older than 12 years only.**

| District / sub-district | Population total | Pre-BA.1 sero-survey | | Post-BA.1 sero-survey | |
| --- | --- | --- | --- | --- | --- |
|  |  | **Number sampled** | **Seroprevalence^1^**  **n (%; 95% CI^2^)** | **Number sampled** | **Seroprevalence^1^**  **n (%; 95% CI^2^)** |
| Anti-S or anti-Gauteng Province | **15,176,113** | 1319 | 1228 (93.1; 91.6-94.3) | 1995 | 1918 (96.1; 95.2-96.9) |
| Johannesburg District | **5,606,238** | 422 | 401 (95; 92.5-96.7) | 732 | 706 (96.4; 94.8-97.6) |
| Johannesburg A | 779,519 | 38 | 36 (94.7; 82.7-98.5) | 140 | 137 (97.9; 93.9-99.3) |
| Johannesburg B | 435,241 | 33 | 32 (97; 84.7-99.5) | 75 | 74 (98.7; 92.8-99.8) |
| Johannesburg C | 799,980 | 73 | 71 (97.3; 90.5-99.2) | 125 | 120 (96; 91.0-98.3) |
| Johannesburg D | 1,396,243 | 113 | 104 (92; 85.6-95.8) | 190 | 181 (95.3; 91.2-97.5) |
| Johannesburg E | 601,433 | 26 | 26 (100; 87.1-100.0) | 37 | 36 (97.3; 86.2-99.5) |
| Johannesburg F | 751,484 | 31 | 30 (96.8; 83.8-99.4) | 46 | 42 (91.3; 79.7-96.6) |
| Johannesburg G | 842,339 | 108 | 102 (94.4; 88.4-97.4) | 119 | 116 (97.5; 92.8-99.1) |
| Ekurhuleni District | **3,825,650** | 393 | 373 (94.9; 92.3-96.7) | 513 | 497 (96.9; 95.0-98.1) |
| Ekurhuleni E1 | 626 517 | 87 | 83 (95.4; 88.8-98.2) | 143 | 140 (97.9; 94.0-99.3) |
| Ekurhuleni E2 | 455 262 | 53 | 51 (96.2; 87.2-99.0) | 62 | 59 (95.2; 86.7-98.3) |
| Ekurhuleni N1 | 708 290 | 75 | 70 (93.3; 85.3-97.1) | 128 | 124 (96.9; 92.2-98.8) |
| Ekurhuleni N2 | 697 175 | 32 | 32 (100; 89.3-100.0) | 37 | 37 (100; 90.6-100.0) |
| Ekurhuleni S1 | 673 758 | 38 | 37 (97.4; 86.5-99.5) | 23 | 22 (95.7; 79.0-99.2) |
| Ekurhuleni S2 | 664 648 | 108 | 100 (92.6; 86.1-96.2) | 120 | 115 (95.8; 90.6-98.2) |
| Sedibeng District | **1,084,503** | 114 | 102 (89.5; 82.5-93.9) | 172 | 168 (97.7; 94.2-99.1) |
| Lesedi | 127 419 | 81 | 74 (91.4; 83.2-95.8) | 136 | 133 (97.8; 93.7-99.2) |
| Midvaal | 126 285 | 20 | 15 (75; 53.1-88.8) | 25 | 24 (96; 80.5-99.3) |
| Emfuleni | 830 798 | 13 | 13 (100; 77.2-100.0) | 11 | 11 (100; 74.1-100.0) |
| City of Tshwane District | **3,709,635** | 232 | 208 (89.7; 85.1-92.9) | 331 | 309 (93.4; 90.1-95.6) |
| Tshwane 1 | 1 032 885 | 94 | 82 (87.2; 79.0-92.5) | 138 | 125 (90.6; 84.5-94.4) |
| Tshwane 2 | 436 950 | 25 | 22 (88; 70.0-95.8) | 47 | 46 (97.9; 88.9-99.6) |
| Tshwane 3 | 730 788 | 52 | 49 (94.2; 84.4-98.0) | 35 | 34 (97.1; 85.5-99.5) |
| Tshwane 4 | 482 448 | 17 | 15 (88.2; 65.7-96.7) | 18 | 17 (94.4; 74.2-99.0) |
| Tshwane 5 | 119 190 | 10 | 9 (90; 59.6-98.2) | 35 | 34 (97.1; 85.5-99.5) |
| Tshwane 6 | 768 446 | 22 | 20 (90.9; 72.2-97.5) | 39 | 35 (89.7; 76.4-95.9) |
| Tshwane 7 | 138 928 | 12 | 11 (91.7; 64.6-98.5) | 19 | 18 (94.7; 75.4-99.1) |
| West Rand District | **950,088** | 158 | 144 (91.1; 85.7-94.6) | 247 | 238 (96.4; 93.2-98.1) |
| Mogale City | 435 254 | 33 | 28 (84.8; 69.1-93.3) | 72 | 69 (95.8; 88.5-98.6) |
| Rand West City | 300 960 | 59 | 58 (98.3; 91.0-99.7) | 95 | 95 (100; 96.1-100.0) |
| Merafong City | 213 874 | 66 | 58 (87.9; 77.9-93.7) | 80 | 74 (92.5; 84.6-96.5) |

^1^Seroprevalence was defined as seropositive for anti-S or anti-N IgG, irrespective of vaccinations status.

^2^CI, confidence interval; Confidence intervals have not been adjusted for multiplicity and should not be used for inference.

**Suppl Table S6: Seroprevalence of SARS-CoV-2 anti-spike (anti-S) or anti-nucleocapsid (anti-N) immunoglobulin G (IgG) Gauteng Province across the Districts and sub-Districts unvaccinated individuals older than 12 years only.**

| District / sub-district | Population total | Pre-BA.1 sero-survey | | Post-BA.1 sero-survey | |
| --- | --- | --- | --- | --- | --- |
|  |  | **Number sampled** | **Seroprevalence^1^**  **n (%; 95% CI^2^)** | **Number sampled** | **Seroprevalence^1^**  **n (%; 95% CI^2^)** |
| Anti-S or anti-N IgG  Gauteng Province | **15,176,113** | 4938 | 3473 (70.3; 69.0-71.6) | 4891 | 4377 (89.5; 88.6-90.3) |
| Johannesburg District | **5,606,238** | 1904 | 1396 (73.3; 71.3-75.3) | 1785 | 1611 (90.3; 88.8-91.5) |
| Johannesburg A | 779,519 | 283 | 201 (71; 65.5-76.0) | 200 | 171 (85.5; 80.0-89.7) |
| Johannesburg B | 435,241 | 164 | 137 (83.5; 77.1-88.4) | 173 | 164 (94.8; 90.4-97.2) |
| Johannesburg C | 799,980 | 352 | 280 (79.5; 75.0-83.4) | 335 | 320 (95.5; 92.7-97.3) |
| Johannesburg D | 1,396,243 | 500 | 350 (70; 65.8-73.9) | 409 | 349 (85.3; 81.6-88.4) |
| Johannesburg E | 601,433 | 130 | 88 (67.7; 59.2-75.1) | 134 | 124 (92.5; 86.8-95.9) |
| Johannesburg F | 751,484 | 197 | 147 (74.6; 68.1-80.2) | 222 | 201 (90.5; 86.0-93.7) |
| Johannesburg G | 842,339 | 278 | 193 (69.4; 63.8-74.5) | 312 | 282 (90.4; 86.6-93.2) |
| Ekurhuleni District | **3,825,650** | 1234 | 872 (70.7; 68.1-73.1) | 1408 | 1294 (91.9; 90.4-93.2) |
| Ekurhuleni E1 | 626 517 | 230 | 139 (60.4; 54.0-66.5) | 193 | 158 (81.9; 75.8-86.7) |
| Ekurhuleni E2 | 455 262 | 159 | 114 (71.7; 64.2-78.1) | 199 | 189 (95; 91.0-97.2) |
| Ekurhuleni N1 | 708 290 | 240 | 151 (62.9; 56.6-68.8) | 214 | 193 (90.2; 85.5-93.5) |
| Ekurhuleni N2 | 697 175 | 193 | 158 (81.9; 75.8-86.7) | 192 | 186 (96.9; 93.4-98.6) |
| Ekurhuleni S1 | 673 758 | 154 | 121 (78.6; 71.4-84.3) | 194 | 181 (93.3; 88.9-96.0) |
| Ekurhuleni S2 | 664 648 | 258 | 189 (73.3; 67.5-78.3) | 416 | 387 (93; 90.2-95.1) |
| Sedibeng District | **1,084,503** | 330 | 219 (66.4; 61.1-71.2) | 382 | 330 (86.4; 82.6-89.5) |
| Lesedi | 127 419 | 241 | 165 (68.5; 62.3-74.0) | 259 | 225 (86.9; 82.2-90.5) |
| Midvaal | 126 285 | 60 | 34 (56.7; 44.1-68.4) | 72 | 63 (87.5; 77.9-93.3) |
| Emfuleni | 830 798 | 29 | 20 (69; 50.8-82.7) | 51 | 42 (82.4; 69.7-90.4) |
| City of Tshwane District | **3,709,635** | 1054 | 691 (65.6; 62.6-68.4) | 981 | 837 (85.3; 83.0-87.4) |
| Tshwane 1 | 1 032 885 | 302 | 188 (62.3; 56.7-67.5) | 278 | 215 (77.3; 72.1-81.9) |
| Tshwane 2 | 436 950 | 109 | 62 (56.9; 47.5-65.8) | 86 | 69 (80.2; 70.6-87.3) |
| Tshwane 3 | 730 788 | 159 | 116 (73; 65.6-79.3) | 91 | 79 (86.8; 78.4-92.3) |
| Tshwane 4 | 482 448 | 45 | 25 (55.6; 41.2-69.1) | 69 | 60 (87; 77.0-93.0) |
| Tshwane 5 | 119 190 | 184 | 116 (63; 55.9-69.7) | 187 | 167 (89.3; 84.1-93.0) |
| Tshwane 6 | 768 446 | 213 | 153 (71.8; 65.4-77.4) | 211 | 192 (91; 86.4-94.2) |
| Tshwane 7 | 138 928 | 42 | 31 (73.8; 58.9-84.7) | 59 | 55 (93.2; 83.8-97.3) |
| West Rand District | **950,088** | 416 | 295 (70.9; 66.4-75.1) | 335 | 305 (91; 87.5-93.7) |
| Mogale City | 435 254 | 87 | 56 (64.4; 53.9-73.6) | 62 | 54 (87.1; 76.6-93.3) |
| Rand West City | 300 960 | 181 | 133 (73.5; 66.6-79.4) | 131 | 122 (93.1; 87.5-96.3) |
| Merafong City | 213 874 | 148 | 106 (71.6; 63.9-78.3) | 142 | 129 (90.8; 85.0-94.6) |

^1^Seroprevalence was defined as seropositive for anti-S or anti-N IgG, irrespective of vaccinations status.

^2^CI, confidence interval; Confidence intervals have not been adjusted for multiplicity and should not be used for inference.

**Supplementary Table S7: Sero-response to SARS-CoV-2 anti-spike (anti-S) and anti-nucleocapsid (anti-N) immunoglobulin G (IgG) in paired samples from individuals who did not receive any Covid-19 vaccine following the pre-BA.1 sero-survey stratified by district, age-groups and vaccination status at pre-BA.1 sero-survey.**

| District | Overall | | Unvaccinated at pre-BA.1 sero-survey | | Vaccinated at pre-BA.1 sero-survey | |
| --- | --- | --- | --- | --- | --- | --- |
|  | **Anti-N IgG^1^**  **n/N (%; 95% CI^3^)** | **Anti-S IgG^2^**  **n/N (%; 95% CI^3^)** | **Anti-N IgG^1^**  **n/N (%; 95% CI^3^)** | **Anti-S IgG^2^**  **n/N (%; 95% CI^3^)** | **Anti-N IgG^1^**  **n/N (%; 95% CI^3^)** | **Anti-S IgG^2^**  **n/N (%; 95% CI^3^)** |
| Gauteng Province | 697/1548  (45; 42.6-47.5) | 680/1548  (43.9; 41.5-46.4) | 497/1070  (46.4; 43.5-49.4) | 553/1070  (51.7; 48.7-54.7) | 200/478  (41.8; 37.5-46.3) | 127/478  (26.6; 22.8-30.7) |
| Johannesburg District | 251/574  (43.7; 39.7-47.8) | 254/574  (44.3; 40.2-48.3) | 180/401  (44.9; 40.1-49.8) | 200/401  (49.9; 45.0-54.7) | 71/173  (41; 34.0-48.5) | 54/173  (31.2; 24.8-38.5) |
| Ekurhuleni District | 266/529  (50.3; 46.0-54.5) | 245/529  (46.3; 42.1-50.6) | 189/366  (51.6; 46.5-56.7) | 210/366  (57.4; 52.3-62.3) | 77/163  (47.2; 39.7-54.9) | 35/163  (21.5; 15.9-28.4) |
| Sedibeng District | 33/77  (42.9; 32.4-54.0) | 31/77  (40.3; 30.0-51.4) | 22/50  (44; 31.2-57.7) | 23/50  (46; 33.0-59.6) | 11/27  (40.7; 24.5-59.3) | 8/27  (29.6; 15.9-48.5) |
| City of Tshwane District | 106/269  (39.4; 33.8-45.4) | 105/269  (39; 33.4-45.0) | 84/191  (44; 37.1-51.1) | 87/191  (45.5; 38.6-52.6) | 22/78  (28.2; 19.4-39.0) | 18/78  (23.1; 15.1-33.6) |
| West Rand | 41/99  (41.4; 32.2-51.3) | 45/99  (45.5; 36.0-55.2) | 22/62  (35.5; 24.7-47.9) | 33/62 (53.2; 41.0-65.1) | 19/37  (51.4; 35.9-66.6) | 12/37  (32.4; 19.6-48.5) |
| Age-group stratification |  |  |  |  |  |  |
| <12 years | 57/126 (  45.2; 36.8-53.9) | 69/126  (54.8; 46.1-63.2) | 57/126  (45.2; 36.8-53.9) | 69/126  (54.8; 46.1-63.2) |  |  |
| 12 to 17 years | 67/127  (52.8; 44.1-61.2) | 84/127  (66.1; 57.5-73.8) | 63/120  (52.5; 43.6-61.2) | 79/120  (65.8; 57.0-73.7) | 4/7  (57.1; 25.0-84.2) | 5/7  (71.4; 35.9-91.8) |
| 18 to 50 years | 376/836  (45; 41.6-48.4) | 352/836  (42.1; 38.8-45.5) | 285/619  (46; 42.2-50.0) | 303/619  (48.9; 45.0-52.9) | 91/217  (41.9; 35.6-48.6) | 49/217  (22.6; 17.5-28.6) |
| >50 years | 193/450  (42.9; 38.4-47.5) | 170/450  (37.8; 33.4-42.3) | 90/198  (45.5; 38.7-52.4) | 99/198  (50; 43.1-56.9) | 103/252  (40.9; 35.0-47.0) | 71/252  (28.2; 23.0-34.0) |

^1^Seroresponse for anti-N is defined for individuals who were seropositive to either S or N at pre-Omicron sero-survey and seroconverted to N or were seropositive to N at pre-Omicron sero-survey and had a ≥2 fold increase in anti-N titers at post-Omicron BA.1 sero-survey.

^2^Seroresponse for anti-S is defined for individuals who were seropositive to either S or N at pre-Omicron sero-survey and seroconverted to S or were seropositive to S at pre-Omicron sero-survey and had a ≥2 fold increase in anti-S titers at post-Omicron BA.1 sero-survey.

^3^CI, confidence interval; Confidence intervals have not been adjusted for multiplicity and should not be used for inference.

*The criteria used for determining seroresponse is outlined in Supplementary Table S2b.

**Supplementary Table S8: Seroconversion, sero-response and overall serological evidence of SARS-CoV-2 infection between the pre-BA.1 and post-BA.1 sero-surveys in individuals who did not receive any Covid-19 vaccine following the pre-BA.1 sero-survey and stratified by vaccination status at pre-Omicron sero-survey.**

|  | Unvaccinated at pre-BA.1 sero-survey | | | Vaccinated at pre-BA.1 sero-survey | | |
| --- | --- | --- | --- | --- | --- | --- |
| District | **Seroconversion^1^**  **n/ N (%; 95% CI^4^)** | **Seroresponse for anti-N and/or anti-S IgG^2^**  **n/ N (%; 95% CI^4^)** | **Overall serological evidence SARS-CoV-2 infection^3^**  **n/N (%; 95% CI^4^)** | **Seroconversion^1^**  **n/ N (%; 95% CI^4^)** | **Seroresponse for anti-N and/or anti-S IgG^2^**  **n/ N (%; 95% CI^4^)** | **Overall serological evidence SARS-CoV-2 infection^3^**  **n/N (%; 95% CI^4^)** |
| Gauteng Province | 349/468  (74.6; 70.4-78.3) | 681/1070  (63.6; 60.7-66.5) | 1030/1538  (67; 64.6-69.3) | 33/42  (78.6; 64.1-88.3) | 252/478  (52.7; 48.2-57.2) | 285/520  (54.8; 50.5-59.0) |
| Johannesburg District | 116/144  (80.6; 73.3-86.2) | 254/401  (63.3; 58.5-67.9) | 370/545  (67.9; 63.9-71.7) | 8/10  (80; 49.0-94.3) | 97/173  (56.1; 48.6-63.3) | 105/183  (57.4; 50.1-64.3) |
| Ekurhuleni District | 124/157  (79; 72.0-84.6) | 255/366  (69.7; 64.8-74.2) | 379/523  (72.5; 68.5-76.1) | 9/10  (90; 59.6-98.2) | 89/163  (54.6; 46.9-62.1) | 98/173  (56.6; 49.2-63.8) |
| Sedibeng District | 20/28  (71.4; 52.9-84.7) | 31/50  (62; 48.2-74.1) | 51/78  (65.4; 54.3-75.0) | 1/2  (50; 9.5-90.5) | 13/27  (48.1; 30.7-66.0) | 14/29  (48.3; 31.4-65.6) |
| City of Tshwane District | 61/101  (60.4; 50.6-69.4) | 107/191  (56; 48.9-62.9) | 168/292  (57.5; 51.8-63.1) | 11/16  (68.8; 44.4-85.8) | 30/78  (38.5; 28.4-49.6) | 41/94  (43.6; 34.0-53.7) |
| West Rand | 28/38  (73.7; 58.0-85.0) | 34/62  (54.8; 42.5-66.6) | 62/100  (62; 52.2-70.9) | 4/4  (100; 51.0-100.0) | 23/37  (62.2; 46.1-75.9) | 27/41  (65.9; 50.5-78.4) |
| Age-group stratification |  |  |  |  |  |  |
| <12 years | 53/74  (71.6; 60.5-80.6) | 82/126  (65.1; 56.4-72.8) | 135/200  (67.5; 60.7-73.6) |  |  |  |
| 12 to 17 years | 30/33  (90.9; 76.4-96.9) | 88/120  (73.3; 64.8-80.4) | 118/153  (77.1; 69.9-83.1) | - | 6/7  (85.7; 48.7-97.4) | 6/7  (85.7; 48.7-97.4) |
| 18 to 50 years | 191/250  (76.4; 70.8-81.2) | 382/619  (61.7; 57.8-65.5) | 573/869  (65.9; 62.7-69.0) | 19/20  (95; 76.4-99.1) | 113/217  (52.1; 45.4-58.6) | 132/237  (55.7; 49.3-61.9) |
| >50 years | 71/106  (67; 57.6-75.2) | 125/198  (63.1; 56.2-69.5) | 196/304  (64.5; 58.9-69.6) | 14/22  (63.6; 43.0-80.3) | 131/252  (52; 45.8-58.1) | 145/274  (52.9; 47.0-58.7) |

^1^Seroconversion is defined for individuals who were seronegative to both S and N at pre-Omicron sero-survey and seroconverted to either S or N at post-Omicron BA.1 sero-survey.

^2^Seroresponse for anti-N and/or anti-S IgG is defined for individuals who were seropositive to either S or N at pre-Omicron sero-survey and either seroconverted to S, seroconverted to N, were seropositive to N at pre-Omicron sero-survey and had a ≥2 fold increase in anti-N titers at post-Omicron BA.1 sero-survey, or were seropositive to S at pre-Omicron sero-survey and had a ≥2 fold increase in anti-S titers at post-Omicron BA.1 sero-survey. ^3^Overall serological evidence of SARS-CoV-2 infection between in the period between the two surveys when the BA.1 wave occurred was defined as either seroconversion or sero-response for anti-N and/or anti-S IgG. ^4^CI, confidence interval; Confidence intervals have not been adjusted for multiplicity and should not be used for inference. *The criteria used for determining seroconversion and sero-response is outlined in Supplementary Table S2b.

**Supplementary Table S9: Seroconversion, sero-response and overall serological evidence of SARS-CoV-2 infection between the pre-BA.1 and post-BA.1 sero-surveys in individuals who received any Covid-19 vaccine following the pre-BA.1 sero-survey and stratified by vaccination status at pre-BA.1 sero-survey.**

|  | Unvaccinated at pre-BA.1 sero-survey | | | Vaccinated at pre-BA.1 sero-survey | | |
| --- | --- | --- | --- | --- | --- | --- |
| District | **Seroconversion^1^**  **n/ N (%; 95% CI^4^)** | **Seroresponse for anti-N^2^**  **n/ N (%; 95% CI^4^)** | **Overall serological evidence SARS-CoV-2 infection^3^**  **n/N (%; 95% CI^4^)** | **Seroconversion^1^**  **n/ N (%; 95% CI^4^)** | **Seroresponse for anti-N^2^**  **n/ N (%; 95% CI^4^)** | **Overall serological evidence SARS-CoV-2 infection^4^**  **n/N (%; 95% CI^5^)** |
| Gauteng Province | 51/71  (71.8; 60.5-81.0) | 81/201  (40.3; 33.8-47.2) | 132/272  (48.5; 42.7-54.4) | 3/5  (60; 23.1-88.2) | 25/85  (29.4; 20.8-39.8) | 28/90  (31.1; 22.5-41.3) |
| Johannesburg District | 20/25  (80; 60.9-91.1) | 51/113  (45.1; 36.3-54.3) | 71/138  (51.4; 43.2-59.6) | 0/1  (0; 0.0-79.3) | 7/25  (28; 14.3-47.6) | 7/26  (26.9; 13.7-46.1) |
| Ekurhuleni District | 12/20  (60; 38.7-78.1) | 15/45  (33.3; 21.4-47.9) | 27/65  (41.5; 30.4-53.7) | 1/1  (100; 20.7-100.0) | 4/23  (17.4; 7.0-37.1) | 5/24  (20.8; 9.2-40.5) |
| Sedibeng District | 2/2  (100; 34.2-100.0) | 3/8  (37.5; 13.7-69.4) | 5/10  (50; 23.7-76.3) | 0/1  (0; 0.0-79.3) | 3/8  (37.5; 13.7-69.4) | 3/9  (33.3; 12.1-64.6) |
| City of Tshwane District | 8/14  (57.1; 32.6-78.6) | 7/18  (38.9; 20.3-61.4) | 15/32  (46.9; 30.9-63.6) | - | 5/13  (38.5; 17.7-64.5) | 5/13  (38.5; 17.7-64.5) |
| West Rand | 9/10  (90; 59.6-98.2) | 5/17  (29.4; 13.3-53.1) | 14/27  (51.9; 34.0-69.3) | 2/2  (100; 34.2-100.0) | 6/16  (37.5; 18.5-61.4) | 8/18  (44.4; 24.6-66.3) |
| Age-group stratification |  |  |  |  |  |  |
| 12 to 17 years | 10/13  (76.9; 49.7-91.8) | 5/18  (27.8; 12.5-50.9) | 15/31  (48.4; 32.0-65.2) | 1/1  (100; 20.7-100.0) | - | 1/1  (100; 20.7-100.0) |
| 18 to 50 years | 34/47  (72.3; 58.2-83.1) | 50/118  (42.4; 33.8-51.4) | 84/165  (50.9; 43.3-58.4) | 0/1  (0; 0.0-79.3) | 10/42  (23.8; 13.5-38.5) | 10/43  (23.3; 13.2-37.7) |
| >50 years | 7/11  (63.6; 35.4-84.8) | 26/65  (40; 29.0-52.1) | 33/76  (43.4; 32.9-54.6) | 2/3  (66.7; 20.8-93.9) | 15/43  (34.9; 22.4-49.8) | 17/46  (37; 24.5-51.4) |

^1^Seroconversion is defined for individuals who were seronegative to both S and N at pre-Omicron sero-survey and seroconverted to N at post-Omicron BA.1 sero-survey.

^2^Seroresponse for anti-N is defined for individuals who were seropositive to either S or N at pre-Omicron sero-survey and seroconverted to N or were seropositive to N at pre-Omicron sero-survey and had a ≥2 fold increase in anti-N titers at post-Omicron BA.1 sero-survey.

^3^Overall serological evidence of SARS-CoV-2 infection between in the period between the two surveys when the BA.1 wave occurred was defined as either seroconversion or seroresponse for anti-N.

^4^CI, confidence interval; Confidence intervals have not been adjusted for multiplicity and should not be used for inference.

*The criteria used for determining seroconversion and seroresponse is outlined in Supplementary Table S2b.

**Supplementary Table S10: Cumulative reported Covid-19 cases, hospitalizations, recorded deaths, and excess mortality in Gauteng Province by age-group and Covid-19 wave**

| Outcomes | Pre-BA.1 period cumulative | BA.1 dominant wave | BA4/5 dominant wave | TOTAL |
| --- | --- | --- | --- | --- |
| Period of case wave | March 3, 2020 Nov. 10, 2021 | Nov. 11, 2021 – April 9, 2022 | April 10, 2022 – June 5, 2022 |  |
| Less than 12 years: Inferred number of infections^1^ | 1,712,830 | 2,057,225 | Not applicable | |
| Recorded cases -no† | 33961 | 21100 | 8149 | 63210 |
| Cumulative case rate per 100,000 population | 1114.3 | 692.3 | 267.4 | 2074.0 |
| Proportion of total cumulative cases, % | 53.7 | 33.4 | 12.9 | 100 |
| Inferred infections: Recorded cases (95%CI) | 50 (47-54) | 98 (88-106) | Not applicable |  |
| Hospitalizations– no.^‡^ | 3832.0 | 2892.0 | 1079 | 7803 |
| Cumulative hospitalisation rate per 100,000 population | 125.7 | 94.9 | 35.4 | 256.0 |
| Proportion of total cumulative cases, % | 49.1 | 37.1 | 13.8 | 100 |
| Inferred infections: Recorded Covid-19 hospitalizations ratio (95%CI) | 447 (418-475) | 711 (640-776) | Not applicable |  |
| Recorded deaths in wave – no. | 135 | 33 | 1 | 169 |
| Cumulative recorded death rate per 100,000 population | 4.4 | 1.1 | 0.0 | 5.5 |
| Proportion of total cumulative cases, % | 79.9 | 19.5 | 0.6 | 100 |
| Inferred infections: recorded Covid-19 deaths ratio  (95%CI) | 12,688 (11,875-13,478) | 62,340 (56,060-67,974) | Not applicable |  |
| Infection Fatality Risk, % | 0.008 | 0.002 |  |  |
| 12-17 years age: : Inferred number of infections^1^ | 955,108 | 1,005,863 | Not applicable |  |
| Recorded cases -no† | 40720 | 19644 | 5911 | 66275 |
| Cumulative case rate per 100,000 population | 3121.2 | 1505.7 | 453.1 | 5080.0 |
| Proportion of total cumulative cases, % | 61.4 | 29.6 | 8.9 | 100 |
| Inferred infections: Recorded cases ratio (95%CI) | 24 (23-25) | 51 (46-55) | Not applicable |  |
| Hospitalizations– no.^‡^ | 1407.0 | 787.0 | 202 | 2396 |
| Cumulative hospitalisation rate per 100,000 population | 107.8 | 60.3 | 15.5 | 183.7 |
| Proportion of total cumulative cases, % | 58.7 | 32.8 | 8.4 | 100 |
| Inferred infections: Recorded Covid-19 Hospitalization ratio  (95%CI) | 679 (646-709) | 1278 (1159-1378) | Not applicable |  |
| Recorded deaths in wave – no. | 48 | 14 | 1 | 63 |
| Cumulative recorded death rate per 100,000 population | 3.7 | 1.1 | 0.1 | 4.8 |
| Proportion of total cumulative cases, % | 76.2 | 22.2 | 1.6 | 100 |
| Imputed infections: recorded Covid-19 deaths ratio (95%CI) | 19,898 (18,927-20,788) | 71847 (65138-77439) | Not applicable |  |
| Infection Fatality Risk, % | 0.005 | 0.001 |  |  |
| 18-50 years age: : Inferred number of infections^1^ | 4,824,076 | 5,881,915 | Not applicable |  |
| Recorded cases -no† | 597308 | 188193 | 56063 | 841564 |
| Cumulative case rate per 100,000 population | 6692.1 | 2108.5 | 628.1 | 9428.7 |
| Proportion of total cumulative cases, % | 71.0 | 22.4 | 6.7 | 100 |
| Inferred infections: Recorded cases ratio(95%CI) | 26 (25-26) | 10 (10-11) | Not applicable |  |
| Hospitalizations– no.^‡^ | 51477.0 | 11094.0 | 2584 | 65155 |
| Cumulative hospitalisation rate per 100,000 population | 576.7 | 124.3 | 29.0 | 730.0 |
| Proportion of total cumulative cases, % | 79.0 | 17.0 | 4.0 | 100 |
| Inferred infections: Recorded Covid-19 Hospitalization ratio  (95%CI) | 94 (92-95) | 530 (504-555) | Not applicable | |
| Recorded deaths in wave – no. | 5666 | 549 | 56 | 6271 |
| Cumulative recorded death rate per 100,000 population | 63.5 | 6.2 | 0.6 | 70.3 |
| Proportion of total cumulative cases, % | 90.4 | 8.8 | 0.9 | 100 |
| Inferred infections: recorded Covid-19 deaths ratio (95%CI) | 8787 (8620-8942) | 1038 (988-1087) | Not applicable | |
| Infection Fatality Risk, % | 0.12 | 0.01 |  | |
| >50 years age: Inferred number of infections^1^ | 838,649 | 1,633,469 | Not applicable | |
| Recorded cases -no† | 255599 | 59355 | 26494 | 341448 |
| Cumulative case rate per 100,000 population | 10092.7 | 2343.7 | 1046.2 | 13482.6 |
| Proportion of total cumulative cases, % | 74.9 | 17.4 | 7.8 | 100 |
| Inferred infections: Recorded cases ratio (95%CI) | 3.3 (3.2-3.4) | 28 (25-30) | Not applicable | |
| Hospitalizations– no.^‡^ | 71400.0 | 7808.0 | 2897 | 82105 |
| Cumulative hospitalisation rate per 100,000 population | 2819.3 | 308.3 | 114.4 | 3242.0 |
| Proportion of total cumulative cases, % | 87.0 | 9.5 | 3.5 | 100 |
| Inferred infections: Recorded Covid-19 Hospitalization ratio (95%CI) | 12.7 (11.4-12.0) | 209 (191-226) | Not applicable | |
| Recorded deaths in wave – no. | 22298 | 1267 | 195 | 23760 |
| Cumulative recorded death rate per 100,000 population | 880.5 | 50.0 | 7.7 | 938.2 |
| Proportion of total cumulative cases, % | 93.8 | 5.3 | 0.8 | 100 |
| Inferred infections: recorded Covid-19 deaths ratio (95%CI) | 38 (37-39) | 1289 (1177-1391) | Not applicable | |
| Infection Fatality Risk, % | 2.66 | 0.08 |  | |

For Covid-19 cases the waves periods for Pre-omicron cumulative, Omicron BA.1 Wave and BA4/5 resurgence were March 3, 2020 to Nov. 10, 2021; Nov. 11, 2021 – April 9, 2022 and April 10, 2022 – June 5, 2022, respectively. For Covid-19 hospitalizations the wave periods for Pre-omicron cumulative, Omicron BA.1 Wave and BA4/5 resurgence were March 7, 2020 to Nov. 21, 2021; Nov. 22, 2021 to April 12, 2022	and April 13, 2022 to June 5, 2022, respectively. For Covid-19 recorded deaths the wave periods for Pre-omicron cumulative, Omicron BA.1 Wave and BA4/5 resurgence were March 31, 2020 to Nov. 23, 2021; Nov. 24, 2021 to May 4, 2022 and May 5, 2022 - June 5, 2022, respectively.
^1^The inferred number of infections in the population pre-Omicron BA.1 was derived by multiplying the seroprevalence in unvaccinated individuals at the time of the pre-Omicron BA.1 sero-survey by the STATS-SA population ^2^^2^ . For post-Omicron BA.1 inferred number of infections was obtained by multiplying the proportion of unvaccinated individuals showing evidence of overall serological evidence of SARS-CoV-2 infection (Table S8) between the pre-Omicron and post-Omicron sero-surveys, by the STATS-SA population. ^2^ The Infection Fatality Ratio was calculated as the inverse of the inferred SARS-CoV-2 infections:recorded deaths or excess ratios.

**Supplementary Figure S1: Seroprevalence in pre- and post-Omicron serosurvey in unvaccinated individuals**

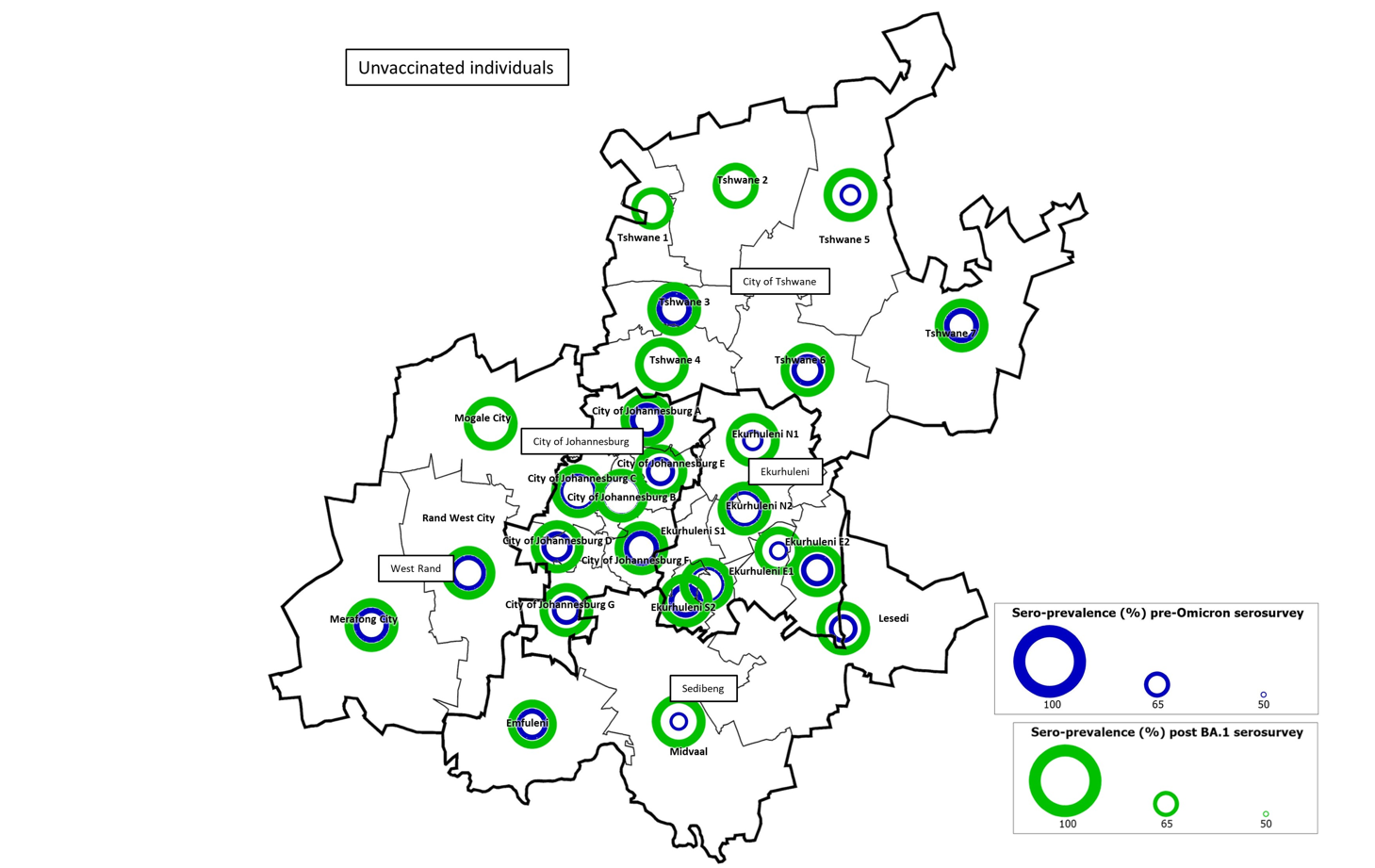

We illustrate the change in seroprevalence pre- and post-Omicron BA.1 in unvaccinated individuals across the 26 subdistricts in Gauteng province
